# Symptom and Age Dependent Casual Effects of Body Size on Bipolar: A Mendelian Randomisation Study

**DOI:** 10.64898/2026.03.25.26349191

**Authors:** Alex Monson, Grace Power, Claire M.A. Haworth, Robyn E. Wootton

## Abstract

**Background:** Previous evidence suggests that higher body size is associated with bipolar disorders, however, whether this association is causal remains uncertain. Interpretation is further complicated by heterogeneity across age, variation in clinical presentation, and potentially distinct underlying aetiologies.

**Aims:** To determine whether body size exerts heterogenous causal effects on bipolar disorder subtypes and symptom profiles.

**Methods:** By leveraging genetic instruments that differentiate effects at different life stages, summary–level univariable and multivariable Mendelian randomisation (MR) analyses were used to estimate how age–specific body size relates to adult psychiatric and symptomatic bipolar features; major depressive disorder (MDD), depressive symptom scores, subthreshold mania symptoms, bipolar disorder, bipolar type I and bipolar type II. Genetic instruments derived from genome–wide association studies (GWASs) for adult body mass index (BMI) (n= 681,275), childhood body size (n= 453,169) and mid-to-later life body size (n= 453,169) served as proxies for prepubertal and adult BMI measures.

**Results:** In univariable MR, higher genetically proxied adult BMI increased the odds of MDD (odds ratio (OR) = 1.13, 95% CI 1.09-1.16), subthreshold mania (OR = 1.09, 95% CI 1.0-1.19)), and depressive scores (β = 0.07, 95% CI 0.05-0.09). There was little evidence that childhood body size had an effect on any outcome. Robust evidence suggested bipolar disorder and MDD increased adult BMI in our reverse univariable analyses.

Using multivariable MR, robust evidence indicated that increased adult body size after accounting for childhood body size increased the odds of MDD, subthreshold mania and depressive scores.

**Conclusions:** Body size may exert different causal effects on bipolar disorder depending on age and symptoms, with detrimental effects occurring during adulthood. Weaker evidence suggested varying effects across bipolar subtypes. Triangulation of findings and higher powered GWASs to detect symptom–specific genetic variants are required to explore whether body size contributes to distinct aetiologies across bipolar patients, informing the identification of novel and personalised treatment targets.

---

Bipolar disorder is characterised by two main affective components of mania and depression.(1) Mania includes increased energy, expansive, elevated or irritable mood and goal–seeking.(2) Depression is defined as sadness, emptiness or hopelessness alongside other symptoms including anhedonia and fatigue.(3) Bipolar disorder comprises two main subtypes: bipolar I (BD–I) and Bipolar II (BD–II). Briefly, BD–I is defined by manic episodes (usually with depressive episodes) whilst BD–II is defined by hypomanic and major depressive episodes according to the DSM–V.(3) Individuals with bipolar disorder experience health, psychosocial and economic impacts such as increased mortality, poorer quality of life and functional impairments.(4) Treatment resistance is common in individuals with bipolar disorder, with non–responses to current medications present in up to one third of patients.(5)

Since life expectancy in patients may be approximately 13 years lower than in the general population,(6) there is a pressing need for more effective treatments. Further understanding of bipolar’s aetiology can identify novel treatments which target specific mechanisms, with aetiologically defined subgroups being used to predict what treatments are most likely to be effective in specific patients, moving towards personalised treatments.(7)

Bipolar disorder is associated with increased morbidity, including impacts on metabolic health such as obesity.(8–10) Associations with obesity are likely bidirectional with possible mechanisms including dysfunctions to hormonal and immunological systems, psychosocial factors, shared genetic liabilities and the effects of psychotropic drugs on weight gain.(11) However, current causal evidence is inconsistent, with modest effect sizes particularly in the direction of obesity on bipolar disorder. One reason for these inconsistencies may be that the effects of body size are often not assessed within specific bipolar subtypes or symptoms for which aetiology may be distinct.(12)

First, there is evidence for a distinct genetic architecture across BD–I and BD–II subtypes.(13,14) However, despite associations between higher body mass index (BMI) and bipolar disorder overall(15), evidence for BMI having differential effects on the main bipolar subtypes is more inconsistent.(16–18) Second, with regards to key bipolar symptoms, higher BMI has more consistent associations with specifically depressive features compared to manic symptoms suggesting different underlying mechanisms.(19,20) Third, another relevant factor that could address inconsistencies in the effects of BMI on bipolar disorder is age. Higher body size in early life may affect hormonal and metabolic processes during puberty which may affect the neurodevelopmental processes involved in the onset of bipolar disorder. Weight stigma may also increase psychosocial stresses particularly during this sensitive period leading to an increased risk when compared to adult body size measures.(21–24) Using data which precedes this sensitive period—such as childhood body size—may be contrasted with adult measures to identify age–dependent effects.

A challenge in epidemiology is ascertaining whether observed effects between risk factors and outcomes are causal, or whether these associations are explained by third–variable confounders, including confounding by undiagnosed existing disease and disease processes (reverse causation).(25,26) Establishing causal effects improves the identification of effective targets for intervention and prevention, by increasing understanding of the underlying components of complex systems involved in pathologies.(27) Over the past two decades, genome–wide association studies (GWASs) have identified increasing numbers of genetic variants—such as single nucleotide polymorphisms (SNPs)—associated with traits of interest.(28) Mendelian randomisation (MR) is a method which attempts to reduce bias from confounding by using these genetic variants as proxies for exposures to estimate their combined effects on an outcome.(29,30) A causal effect between the exposure(s) and outcome can be inferred if various core conditions are met (outlined in Methods). Multivariable MR (MVMR) extends the standard framework by incorporating multiple exposures, enabling the adjustment of SNP–exposure effects for potential confounding pathways or mediation pathways.(31,32) In a lifecourse framework, MVMR allows the effects of childhood and adult body size to be estimated independently of each other, thereby isolating age–specific effects that may otherwise be conflated when using univariable MR approaches.(33)

In this paper, we aim to first explore the heterogeneity amongst bipolar phenotypes by assessing the genetic correlations between traits to clarify the underlying shared liabilities across their different bipolar features, related conditions, and their shared liabilities with measures of body size. We then aim to account for possible heterogeneity using MR approaches by disentangling effects of body size on: 1) diagnoses of major depressive disorder (MDD), bipolar disorder (encompassing both BD–I and BD–II) 2) and specific bipolar subtypes (BD–I and BD–II separately), 3) specific bipolar symptoms (subthreshold mania and depression), and 4) at different ages (childhood v. adulthood). We assess possible reverse causation by using MDD or bipolar disorder as exposures on adult BMI. We assess MDD in addition to bipolar disorder, which allows us to explore how body size aetiology may vary across a spectrum of depression and bipolar features, especially as genetic overlap with MDD varies across bipolar subgroups, suggesting heterogenous mechanisms.(13)

## METHODS

This study follows the Strengthening The Reporting of Observational Studies in Epidemiology Using Mendelian Randomisation (STROBE-MR) guidelines (provided in the Supplement)(34). The study protocol was not preregistered.

We adopt a summary–level MR design(35) which uses two genome–wide association studies (GWASs) for the exposure and the outcome.(28) The causal interpretation from this analysis relies on certain core assumptions being met. The first (IV1) is the ‘relevance’ assumption, which states that the instrument Z (SNPs) must be robustly associated with the exposure. The second (IV2), is the ‘independence’ assumption, which states that the Z–Y association should be independent of genetic confounds which may be induced through population stratification, assortative mating or dynastic effects. The third assumption (IV3), ‘exclusion restriction’, states that instruments should only act on the outcome (Z–Y) through the exposure (X). IV3 violations may occur due to ‘horizontal pleiotropy’ in which a single variant may have multiple biological effects which act on Y via pathways that do not go through X (See Figure 1).(29)

**Figure 1.**
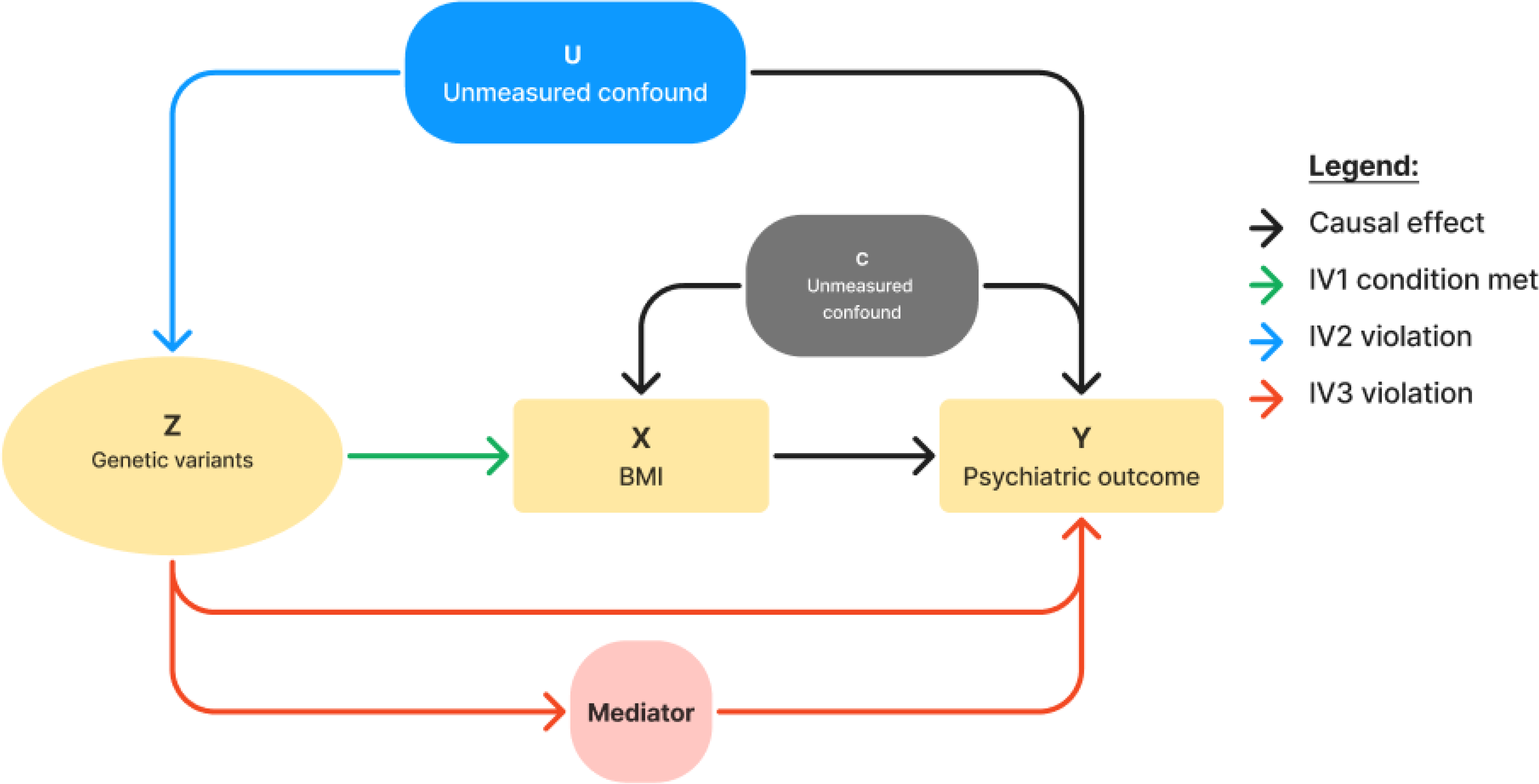
Directed Acyclic Graph (DAG) Displaying the Main Mendelian Randomisation Core Conditions in the Context of the Forward Univariable Analyses *Note.* Z indicates genetic variants in the form of multiple SNPs associated with the exposure X (body size). Y indicates the outcome e.g., a diagnosis of MDD or bipolar. C indicates a confounder of the X−Y relationship, of which Z is independent of. U indicates a confound of the Z−Y relationship. The green line indicates the IV1 condition violation of ‘relevance’ states Z must be robustly associated with X. The blue line indicates the IV2 condition violation of ‘independence’ states the Z−Y relationship must be independent of confounding. The red line indicates the IV3 condition violation of ‘exclusion restriction’ states that Z must only be associated with Y through X.

### Overview of data sources

All data sources used full summary statistics from mixed–sex participants of European ancestry to maintain maximum power whilst avoiding confounding through population stratification, reducing the risk of violating the second IV condition.(36) We used the largest GWASs available to maximise statistical power. Post–hoc power calculations are reported in Supplementary Table 12. Adjustment for covariates differed across GWASs but included age, sex, and genetic principal components; please see the individual GWAS papers for further details (37–46). As exposure and outcome samples are derived from ancestrally homogenous populations with similar controls for population structures applied, the SNP–exposure associations are applicable to the outcome samples. Unless specifically stated, age was not consistently reported across data sources but GWASs recruited participants across age ranges, though typically during mid to late adulthood. Thus, we interpret the adult BMI and body size alongside the psychiatric/symptom GWASs to estimate lifetime effects, however caution should be taken in assuming that genetic effects are invariant across age.(47) A summary of our data sources is reported in Supplementary Table 1.

### Summary statistics for body size

*Adult BMI*. For Adult BMI we obtained full summary statistics from the GWAS by Yengo et al. (2018)(40) of 681,275 adult participants, recruited from the UK Biobank (UKB)(48) and a previous GWAS by Locke et al. (2015).(49) BMI was measured or self–reported as participants weight divided by height squared (kg/m^2^). Although the mean age of the complete sample was not reported, participants across studies were recruited at all adult life stages, though typically sampled from mid to late adulthood.

*Childhood body size* was obtained from a GWAS conducted by Richarson et al. (2020)(44) using 453,169 UKB participants (data Data–Field 1687: https://biobank.ctsu.ox.ac.uk/crystal/field.cgi?id=1687) (50,51) and retrospectively asked “When you were 10 years old, compared to average would you describe yourself as thinner, about average, or plumper?” Effect estimates can be interpreted as the change in outcome odds per 1 SD increase in liability to body size reflecting increased likelihoods of being classified into higher weight categories; from ‘thinner’ to ‘about average’ and from ‘about average’ to ‘plumper’.(52,53) Analyses were conducted using a linear regression model assuming that genetic instruments incur equal effect between each comparative weight category.(44)

*Adult body size.* An adult body size variable was derived using clinically measured BMI data from 453,169 UKB participants (mean age 56.5 years). It was then separated into a 3-tier variable using the same categories as the childhood body size measure for comparability; “thinner” (21.1 kg/m^2^-25 kg/m^2^), “about average” (25 kg/m2-31.7 kg/m^2^) and “plumper” (31.7 kg/m^2^-59.9 kg/m^2^). These scores have been independently validated in three different cohorts, providing verification that these genetic instruments can reliably separate childhood and adult BMI.(44–46) Childhood and adulthood body size were used as proxies for age–specific BMI.

### Psychiatric summary statistics

As large powered GWASs often use the same cohorts such as the UKB, we selected subsets of outcome GWASs with UKB participants removed for MDD and bipolar disorder to mitigate against biases that may arise from sample overlap in summary–level MR (54). We also used subsets of the MDD and bipolar disorder outcome datasets with 23andme data removed as this company does not allow for access to their full summary statistics without special permission.(55)

*MDD.* GWAS data for 357,636 MDD cases and 1,281,936 controls was obtained from the newest MDD GWAS dataset published by the Psychiatrics Genomics Consortium (PGC) across 74 clinical and population samples, defining ever reporting MDD cases via medical records, clinical diagnostics and interviews, questionnaires and other self–report measures. Diagnostic criteria included the ICD–9, ICD–10, DSM–IV, or DSM-5 classifications for major depression.(37) We also used MDD as an exposure in our reverse univariate MR analyses, which includes full data from the UKB and 23andme. We derive our instruments from data providing the top 10,000 independent SNP–exposure associations using 525,197 cases and 3,362,335 controls.

*Bipolar disorder.* Our bipolar disorder outcome data was derived from the newest PGC bipolar disorder GWAS(38) across 69 cohorts of 57,833 cases and 722,909 controls. For our outcome bipolar disorder measure, we used a subset with UKB and 23andme samples removed. Bipolar disorder was defined through diagnostic interviews, questionnaires, medical records, registry data and self–report data using clinical and community samples. Diagnostics were made using ICD–9, ICD–10, DSM–III, DSM–IV, DSM–V or rapid diagnostic centre classifications and included both BD–I and BD–II patients. We used bipolar disorder as an exposure in our reverse analyses, again including full data from the UKB and 23andme, using the top independent SNP–exposure associations, identified from the derivative GWAS, provided from 131,969 cases and 2,322,416 controls.

*Bipolar subtypes.* Our psychiatric outcomes are also split by bipolar subtype using the previous PGC bipolar disorder GWAS(39) consisting of 55 cohorts for BD–I (25,060 cases, 449,978 controls) and 31 cohorts for BD–II (6,781 cases, 364,075 controls). Cases meet criteria for lifetime bipolar disorder (via DSM IV or ICD–9 or ICD–10). Diagnoses were established through diagnostic interviews, medical records or clinical checklists. Diagnostics were made using DSM–IV, ICD–9 or ICD–10 classifications.

*Schizophrenia.* Alongside our phenotypes outlined in Supplementary Table 1, we added schizophrenia as a variable to our genetic correlation analyses due to evidence that genetic covariance with schizophrenia may be greater in BD–I compared to BD–II.(56,57) This further contextualises any divergences in covariance with BMI across bipolar subtypes. We used the latest PGC GWAS of schizophrenia using 76 cohorts which includes 52,017 cases of patients diagnosed with schizophrenia or schizoaffective disorder or other schizophrenia spectrum disorders—according to ICD or DSM classifications—and 75,889 controls.(58) Cases were identified through medical records/registers, diagnostic interviews/assessments or mixed strategies.

### Symptom–level summary statistics

*Ever subthreshold manic.* The subthreshold mania symptom instrument was taken from Jiang et al. (2021)(41) using 146,837 (8,449 participants responded ‘yes’) participants from the UKB (data field 4642 – https://biobank.ctsu.ox.ac.uk/crystal/field.cgi?id=4642). Participants were asked “Have you ever had a period of time lasting at least two days when you were feeling so good, "high", excited or "hyper" that other people thought you were not your normal self or you were so "hyper" that you got into trouble?". Responses were restricted to ‘yes’ or ‘no’.(59)

Age was not reported directly, however the UKB has recruited from participants aged between 40 and 69 at enrolment.(50) As this measure is taken from a population sample and does not fulfil the DSM–V criteria for durations (of at least 4 days) or full criteria of symptoms for hypomania, we interpret this measure as a proxy for subthreshold mania.(3) We use a population measure of subthreshold mania to measure manic symptoms, as high powered GWASs for clinical manic symptoms are not currently available.

*Depressive symptom score.* The depressive symptom score GWAS summary statistics were taken from Okbay et al.(2016)(42) which incorporated both self–report and diagnostic data across 3 cohorts from a total of 161,460 participants. UKB(50) depressive scores were calculated by summing and standardising the responses to the following 2 questions: “Over the past two weeks, how often have you felt down, depressed or hopeless?” and “Over the past two weeks, how often have you had little interest or pleasure in doing things?”. Responses ranged from: “Not at all” to “Nearly every day”. Higher scores were further recorded if participants either had two separate depression diagnoses throughout their lifetime, or a diagnosis of MDD (according to DSM–IV or ICD classifications) to create a total depression score.

### Statistical analysis

*Genetic correlations.* We conducted genetic correlation analyses using linkage disequilibrium (LD) score regression (LDSC) through the LDSC Python package version 2.0.1.(60) LDSC estimates the genetic correlations between traits by regressing an estimation of genetic covariance against LD scores.(60) As LD is uncorrelated with external confounding factors like population stratification, LDSC gives a robust estimation of genome–wide genetic correlation.(60)

*Univariable Mendelian Randomisation*. Univariable MR analyses were run to estimate the total effects of the genetically predicted exposures on outcomes of interest, as depicted by the Z–X–Y association in Figure 1. Exposure SNPs were selected from full GWAS summary data based on genome–wide significance (p<5x10^–8^). We select independent SNPs through clumping by removing other SNPs within regions at r^2^ above 0.01 and within 1000 kilobases. The SNPs where then harmonised using the TwoSampleMR package version 0.6.2.(61) in which effect alleles were matched across datasets, with palindromic SNPs with similar allele frequencies (≈50%) removed.(61,62). To explore evidence for reverse causation, we used bipolar disorder and MDD as exposures separately to test their effects on adult BMI as the outcome.

Multivariable *Mendelian Randomisation (MVMR).* To further assess evidence for age–dependent effects we test a framework in which childhood and adult body size respectively may exert independent casual effects on our outcomes (used in the forward univariable analyses) and where adult body size may act as a mediator between childhood body size and our outcomes using MVMR analysis (see Figure 2).(33) We derived the exposures of adult body size adjusted for child body size and child body size adjusted for adult body size. Consequently, we estimate the direct effects between child and adult body size respectively on our outcomes. We obtained an IVW estimate for our effects.(63) Analyses were conducted using the MVMR package version 0.4.(64)

**Figure 2:**
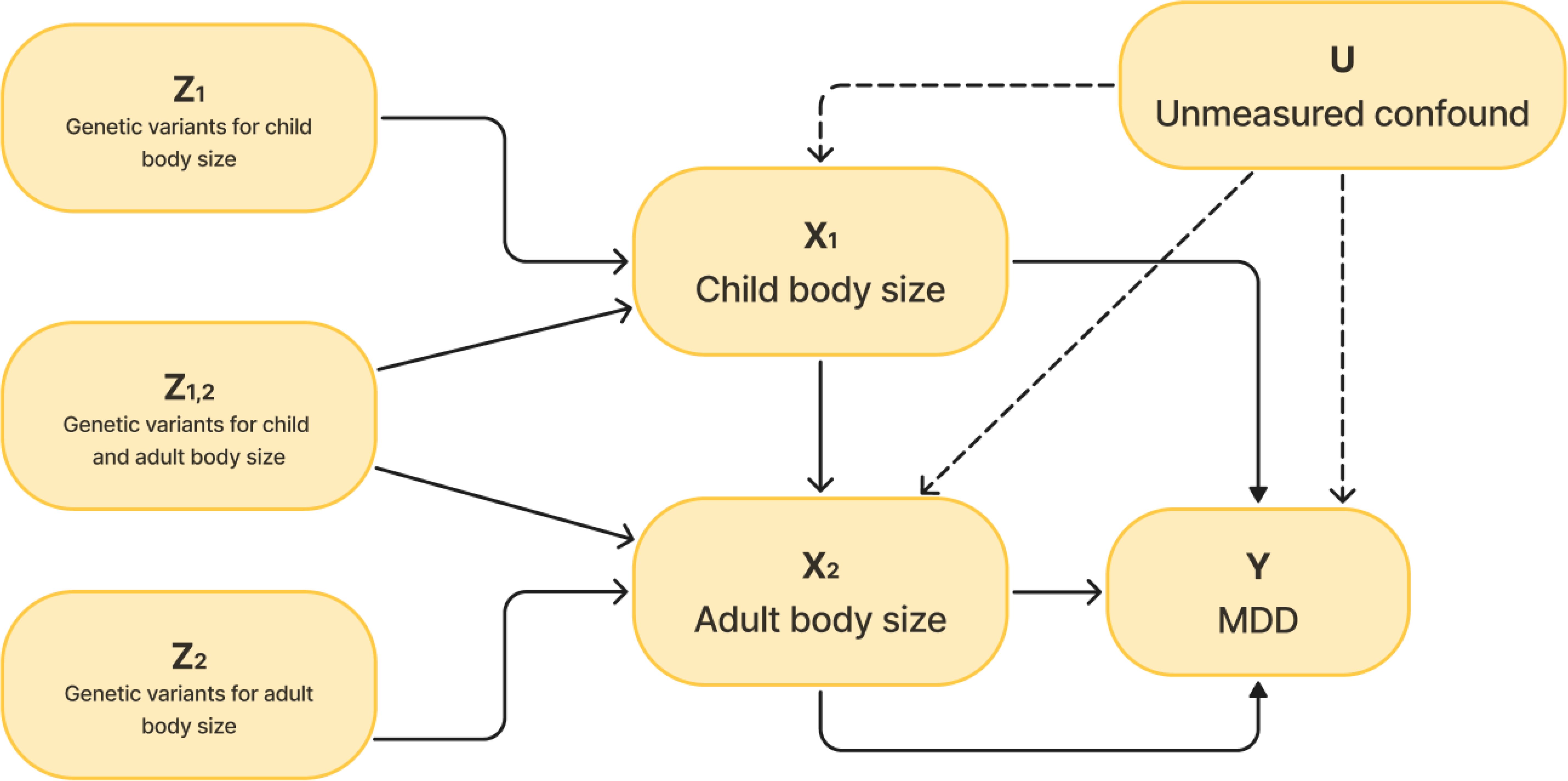
*Directed Acyclic Diagram for Proposed Directions in the Multivariable MR Analysis* *Note.* Z_1_ indicates the genetic variants (IVs) robustly associated with child body size (X_1_) independently of Z_1,2_ whilst Z_2_ indicates the genetic variants robustly associated with adult body size (X_2_) independently of Z_1,2_. Z_1,2_ indicates genetic variants that are robustly associated with both child and body size. Y indicates the outcome and U indicates an unmeasured confound between the outcome and the exposures. The solid arrow between X_1_ or X_2_ respectively and Y indicates the direct effects between each exposure and Y, whilst the line arrows indicate the indirect effect between X_1_ and Y through X_2_ as a mediator.

*Inverse variance weighted (IVW) method.* The inverse variance weighted (IVW) method was the main analysis method used to estimate the effect of the exposures on outcomes of interest in both the univariable and multivariable analyses. IVW uses fixed–effect meta–analyses to take the average effect of the exposure SNPs on the outcome, weighted on the inverse variance, such that SNP estimates with smaller standard errors are given greater weighting in the overall estimate.(35) Statistical analyses were conducted in R version 4.3.2(69).

### Sensitivity analyses to test the IV conditions

The IV1 assumption (relevance) was assessed using F–statistics, or conditional F–statistics (for the multivariable MR analysis).(65) To examine the robustness of the IV3 assumption (no horizontal pleiotropy), we conducted three additional sensitivity methods (MR–Egger, weighted median, weighted mode) which make different assumptions about horizontal pleiotropy. Although violation of the IV3 assumption cannot be directly tested, consistency in the direction of effects across these methods can be regarded as stronger support against bias from horizontal pleiotropy.(29,66) Additionally, heterogeneity statistics and MR Egger intercept tests were analysed to further assess the robustness to bias from IV3 violation. We used Steiger filtering to remove SNPs where reverse causation was evidenced. Each SNP’s variance explained in the exposure and the outcome was calculated, and Steiger filtering was applied if less than 90% of the total SNPs for each exposure had a greater variance explained in the exposure than the outcome. In such cases, SNPs with greater outcome variance explained were removed and the main analyses were rerun.(67) We conducted leave–one–out analyses to assess the effects of removing single SNPs on the overall IVW estimates.(29) Scatter, leave–one–out and single SNP plots were visually inspected as a sensitivity check to identify possible outliers. MR PRESSO(68) effect estimates and tests were used to remove outlying SNPs due to possible horizontal pleiotropy. Further details of the methodology and results of these sensitivity tests are reported in the Supplementary Material.

## RESULTS

### Genetic correlation analyses

Adult BMI was positively correlated with adult and child body size, MDD, subthreshold mania and depressive symptom scores whilst being negatively correlated with schizophrenia (rG = -0.11 95% CI [-0.14, -0.08]) and possibly BD–I (rG = -0.06 95% CI [-0.1, -0.02]). Child body size was only negatively associated with schizophrenia (rG = –0.1, 95% CI [–0.13, –0.06]) and positively with adult body size (rG = 0.54, 95% CI [0.51, 0.58]; Figure 3).

**Figure 3:**
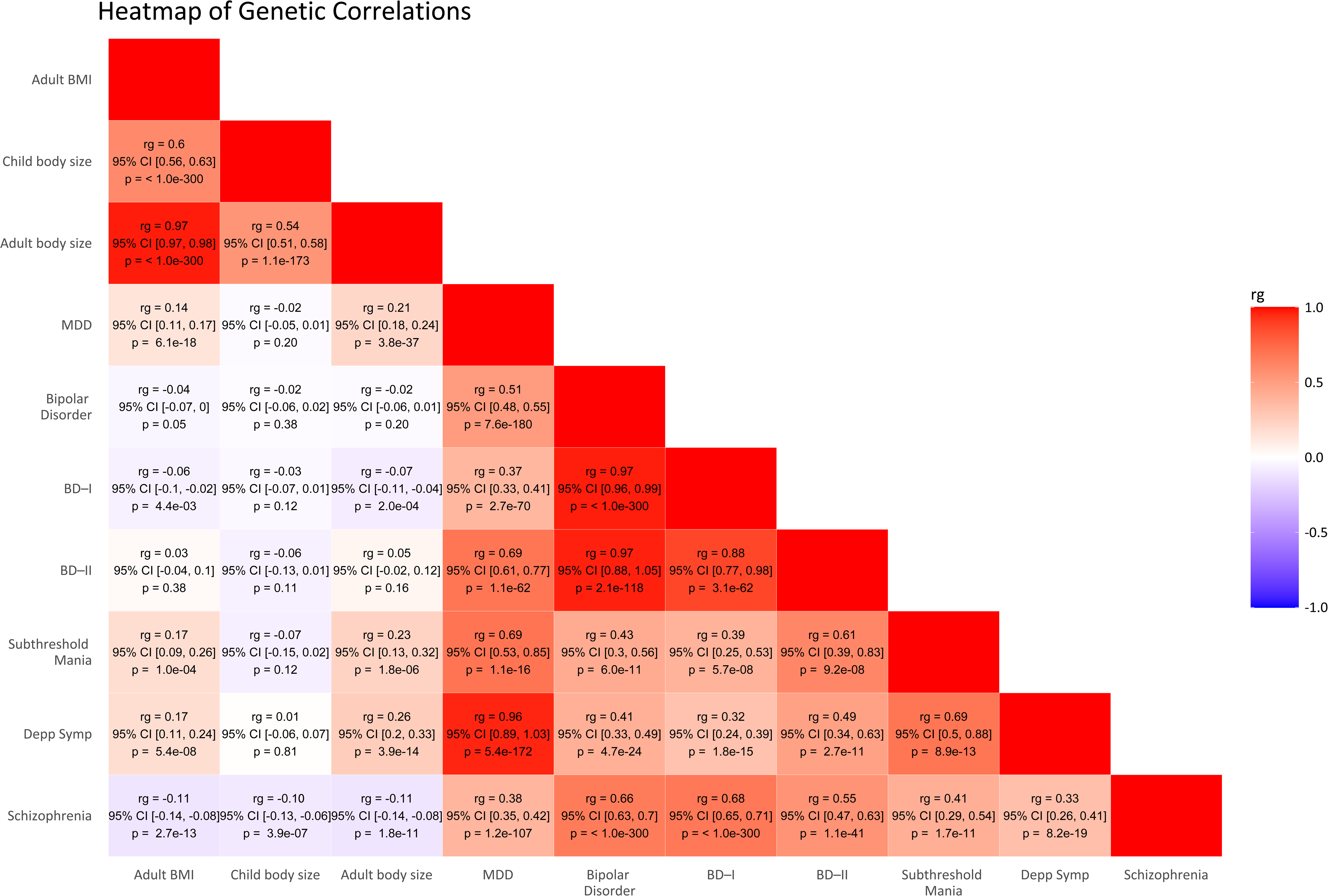
*Heatmap for LDSC Regression of Genetic Correlations* *Note.* ‘rg’ denotes the genetic correlation between variables with their 95% confidence intervals provided. Squares with a redder colour indicate rgs closer to 1 and bluer colours indicate rgs closer to –1. ‘BS’ in child BS and adult BS stands for body size. ‘Depp Symp’ stands for depressive symptom scores. BD, BDI and BDII stand for bipolar disorder, bipolar I and bipolar II respectively. MDD stands for major depressive disorder.

### Mendelian randomisation analyses

*Testing for causal effects of BMI on bipolar features and MDD.* In the univariable MR analysis, (IVW), adult BMI showed robust evidence for increasing the odds of MDD (OR = 1.13 95% CI [1.09, 1.16]) and subthreshold mania (OR = 1.09 95% CI [1.00, 1.19]) as well as increasing depressive symptom scores (β = 0.07 95% CI [0.05, 0.09]). There was little evidence of effects on bipolar disorder, BD–I or BD–II. There was also little evidence of effects of childhood body size on any of the outcomes. Findings are displayed in Figure 4 and Supplementary Table 2. Reproduction of the SNP–level effects can be obtained using the analysis code provided on Github (https://github.com/alex-monson/Age-and-Symptom-dependent-effects-between-body-size-and-bipolar) using publicly available GWAS data.

**Figure 4:**
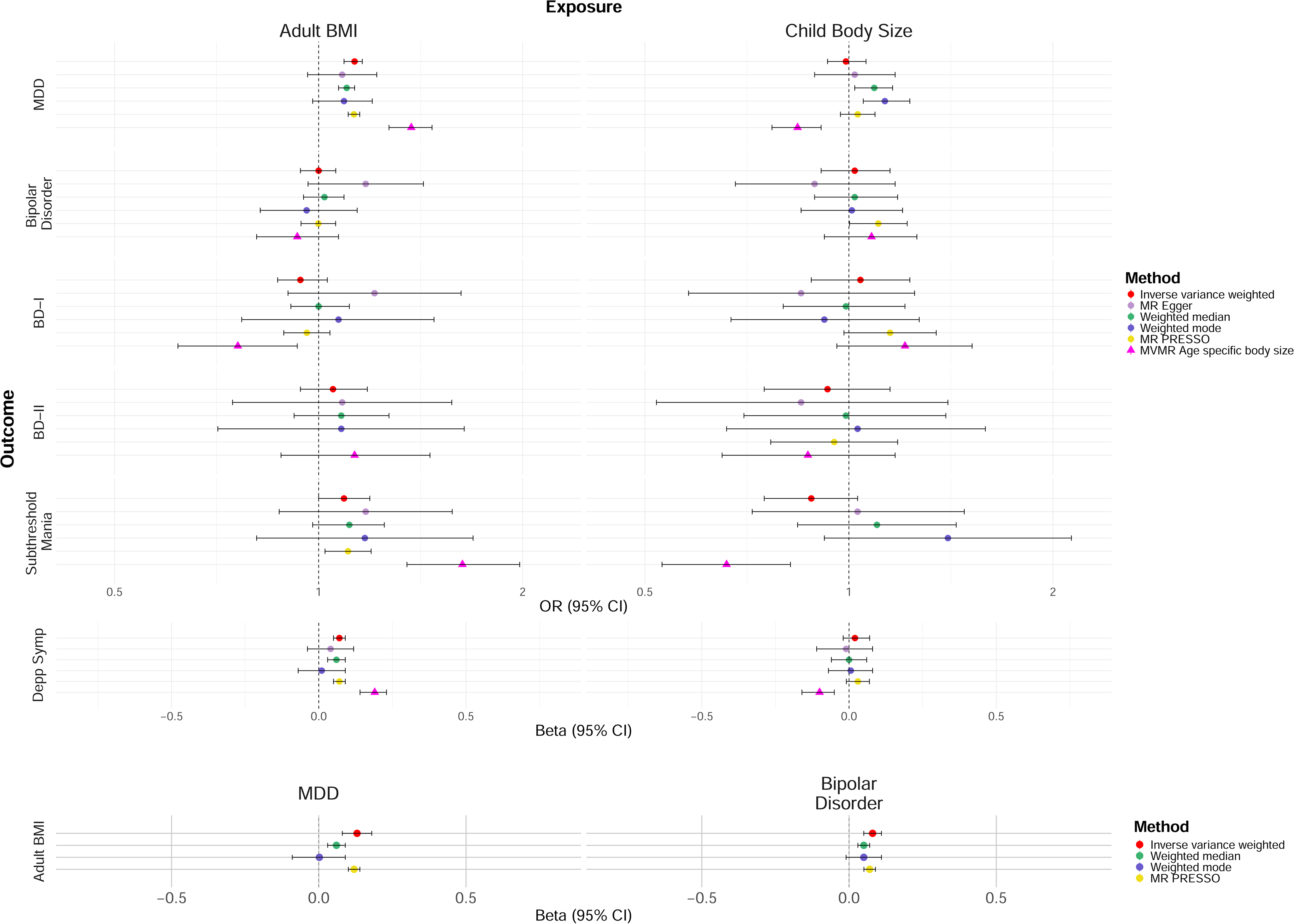
*Forest Plot of Univariate MR Analyses* *Note.* Circular point estimates indicate univariable analyses whilst triangular estimates indicate the multivariable estimates. MR PRESSO is not reported if no outliers were detected. MR Egger is not reported when I^2^ statistics were below 0.6. The odds ratio and beta scales are not proportionate to each other. ^a^Adult BMI effects are estimated as the change in odds of the outcome per 1 SD increase in genetically predicted BMI (measured in kg/m²). ^b^Child body size effects estimate the change in odds of the outcome per 1 SD increase in genetic liability to body size, corresponding to an increased probability of being classified into higher body size categories (e.g., from ‘thinner’ to ‘about average’, or from ‘about average’ to ‘plumper’). MDD stands for major depressive disorder, BD, BDI and BDII stand for bipolar disorder and bipolar type I and II respectively. ‘Depp symp’ denotes depressive symptom scores.

*Testing for the reverse effects of MDD and bipolar disorder on adult BMI.* For our bidrectional analyses, we found robust evidence that having MDD increased adult BMI (β = 0.13, 95% CI [0.08, 0.18]). We also found evidence that bipolar disorder increased adult BMI (β = 0.08, 95% CI [0.05, 0.11]).

*MVMR analysis assessing age–dependent effects of body size.* We found strong evidence suggesting that increased adult body size, after accounting for child body size, increased the odds of MDD (OR = 1.37, 95% CI [1.27 to 1.47, subthreshold mania (OR = 1.63 [1.35 to 1.98]) as well as increasing depressive symptom scores (β = 0.19 [0.14 to 0.23]), while decreasing the odds of BD–I (OR = 0.76 [0.62 to 0.93]). Strong evidence indicated that child body size, after accounting for adult body size, decreased the odds of MDD (OR = 0.84, 95% CI [0.77, 0.91]), subthreshold mania (OR = 0.66 [0.53 to 0.82]), and decreased depressive symptom scores (β = -0.10 [-0.16, -0.05]) (Figure 4). The full results are reported in Supplementary Table 3.

### Sensitivity checks

The relevance condition was met for all exposures, with F–statistics and conditional F–statistics for instruments above 10 (Supplementary Tables 1 and 3). There was no strong evidence for violation of the IV3 condition via MR Egger intercept tests, except for adult BMI on BD–I which suggested weak evidence for possible bias from horizontal pleiotropy (p = .08) (Supplementary Table 5). Leave–one–out plots indicated that removing any SNPs did not affect the conclusions of our analyses (Supplementary Figures 3b-7b). Inspection of scatter plots and single SNP forest plots indicated the presence of outliers across multiple tests (Supplementary Figures 3c-7c). We report outlier–robust MR PRESSO estimates alongside our main results in Supplementary Table 8. The distortion test revealed substantial evidence for outliers affecting the overall effect estimates only for adult BMI on bipolar disorder (p = .03), suggesting a substantial change to the effect estimate from the removal of outliers.(68) However, this did not change our interpretations as both IVW and MR PRESSO methods predicted very weak evidence for this effect. MR PRESSO tests did indicate evidence for childhood body size affecting bipolar disorder (OR = 1.11, p = .05) and BD–I (OR = 1.15, p = 0.06) which deviate from the conclusions of the IVW and sensitivity analyses. Evidence suggested substantive reverse causation (total SNPs where variance explained in the exposure was greater compared to in the outcome was below 90%) for adult BMI on BD–II, child body size on BD–I, BD–II and subthreshold mania and MDD on adult BMI. Analyses after Steiger filtering did not change our conclusions (Supplementary Tables 9-10, Supplementary Figure 1). Q statistics indicated substantial heterogeneity for all tests except for childhood body size on subthreshold mania (p = .07). Further details on the sensitivity checks, including I^2^ statistics and SIMEX corrections are reported in the Supplementary Tables 4-10.

## DISCUSSION

Our study aimed to assess the effects of body size on bipolar features using a Mendelian Randomisation framework as a tool for causal inference. Due to the high heterogeneity of bipolar disorder, we explored subtype–dependent, symptom–dependent and age–dependent factors that may moderate these associations, to identify what features of bipolar disorder body size may act on and at which life stages. This may help determine individuals who are more vulnerable to specific risk factors, such as higher body size, and inform targeted interventions at relevant life stages.(70)

We found that increased adult BMI increased odds for MDD, ever having a subthreshold manic episode and depressive symptom scores but not bipolar disorder, nor its main subtypes, BD–I or BD–II. Our results replicate previous MR findings suggesting a causal effect of higher BMI on MDD.(71,72) However, our analysis extends this work by examining effects on both MDD alongside a continuous depression measure and comparing findings with a range of bipolar related outcomes. Evidence of causal effects of body size on depression are also supported by longitudinal findings of dietary interventions in reducing depressive symptoms in patients with obesity.(73) Explanations linking higher body size to depression include increased social stigma(74) and decreased quality of life(75) alongside biological mechanisms like shared genetic liability and immune–metabolic pathways involving inflammation and hormone dysregulation (e.g., insulin, cortisol and leptin).(76) Thus, effects of body size on depression have been consistently evidenced across clinical and population samples.

Consequently, the lack of evidence linking body size to bipolar disorder is notable, since depression is one major component of the condition. These conclusions challenge observational associations between obesity and bipolar disorder(15), and similarities in proposed mechanisms linking obesity to bipolar disorder and other psychiatric outcomes like MDD and schizophrenia.(77) Although we found little evidence of an effect of BMI on BD–I or BD–II, this ‘subtyping’ approach did reveal weak evidence for effects of higher BMI acting in opposite directions; negatively with BD–I and positively with BD–II. When combined into a single bipolar phenotype, these opposing effects likely offset each other, yielding an overall estimate close to the null. Our genetic correlation analyses provide possible support for this, where BD–I was more strongly genetically correlated with schizophrenia than MDD, but BD–II was more strongly genetically correlated with MDD than schizophrenia. Higher BMI negatively predicted schizophrenia in another MR study,(71) therefore BMI effects may exist on a scale between positive associations with MDD and negative with schizophrenia in which BD–I and BD–II lie in–between (see Supplementary Figure 2). DSM criteria of bipolar subtypes are not designed to differentiate aetiology and are subject to further misclassification biases compared to our age sensitive measures, further explaining why subtyping approaches provide weaker evidence for effects. Overall, higher powered GWASs of aetiologically defined bipolar subtypes are required to further explore this hypothesis. Furthermore, our subthreshold manic outcome increased with higher adult BMI and was found to be highly genetically correlated with MDD and BD–II. This suggests that bipolar features more similar to MDD share positive associations with BMI. Indeed, individuals with BD–II or with depression at greater bipolar risk are more likely to experience atypical depression features; partially characterised by weight gain.(78)

Subthreshold bipolar features are common in MDD patients(79,80) and may explain the high genetic overlap between MDD and the subthreshold mania phenotypes.

In relation to age–dependent total effects assessed using univariable MR, childhood body size showed no clear evidence of an effect on any of the outcomes investigated. This conflicts with observational findings reporting associations between childhood adiposity and adult depression.(81) MVMR analyses further support the age–dependent effects of adult body size, independent of childhood body size, on increased risk of adult MDD, subthreshold mania and higher depressive symptoms. The subthreshold mania outcome showed the same patterns as the depression measures but with larger effect sizes, suggesting that subthreshold manic features could indicate a possible depression subgroup where higher adult body size exerts a greater mechanistic effect. However increased childhood body size, independent of adult body size, indicated a protective effect on these outcomes. This disputes observational findings in the UKB that childhood body size, adjusted for adult BMI, predicts adult depression(82) but corroborates recent MVMR work by Pathak et al. (2025) finding that childhood body size independent of adult body size predicted protective effects on adult MDD.(83) Retrospective comparisons of body size at age 10 may introduce interpretative or recall biases and be qualitatively different than objective and more contemporary body mass measures. Nonetheless, this measure has been validated with current external datasets in predicting childhood obesity,(84) and our genetic correlation analyses found strong relatedness with adult BMI.(44–46) Alternately negative associations may be driven by detrimental effects on depression and subthreshold mania at low body sizes.(85) Our MVMR analyses suggest the benefits of using age–specific measures wherein childhood specific effects appear to oppose adulthood specific effects, which may mask heterogeneous effects if not addressed. Additionally, the MVMR analyses provided some evidence for subtype–specific effects whereby higher adjusted adult body predicted a protective effect on bipolar–I which warrants further investigation.

For our reverse analyses we found that lifetime MDD and bipolar disorder predicted increased adult BMI. As MDD and bipolar disorder onset typically occurs during early adulthood and the BMI GWAS typically recruited participants during mid to late adulthood, it is plausible temporally that these mood disorders may increase body size throughout adulthood.(86,87) Features of atypical depression include weight gain and increased appetite, being components of depression in both MDD and bipolar disorder.(3) Mood disorders also predict lifestyle changes associated with metabolic outcomes.(88) Therefore, our findings support the proposed bidirectional associations between obesity and depression but not bipolar disorder.(12) This may be explained by differences in heterogeneity across depression and bipolar disorder.

### Strengths & limitations

To our knowledge this is the first MR study assessing the effects of body size on bipolar features such as specific symptoms and subtypes. This takes a relatively overlooked approach in the MR context, by assessing body size effects on bipolar disorder whilst explicitly accounting for psychiatric genetic heterogeneity. Where causality is evidenced by the IVW estimate, our sensitivity analyses generally show consistent direction with comparable magnitudes, supporting the robustness to bias from horizontal pleiotropy (IV3 violation). These findings strengthen confidence in our causal estimates, although triangulation with other study designs remains necessary to support stronger causal claims.(89)

There were limitations to our findings. For instance, I^2^ (reported in the Supplementary Materials) for our adult BMI and psychiatric exposures were lower than 0.9. This suggests potential violation of the ‘No Measurement Error’ (NOME) assumption which will bias MR Egger estimates towards the null unless the ‘INstrument Strength Independent of Direct Effect’ (InSIDE) assumption is also violated; in which overall directions of bias may be hard to predict.(90) Generally I^2^ and Q statistics were high indicating high heterogeneity and possible horizontal pleiotropy(91), although none of our MR Egger intercept tests indicated evidence for IV3 violation, providing evidence against consistent confounding via directional horizontal pleiotropy.(92) Also, although our obesity measures include SNPs in regions that have been functionally associated with adiposity such as FTO,(93) high–powered GWASs can detect distally related genetic effects increasing pleiotropy and threatening the gene–environment equivalence assumption which states that a genetic effect on an exposure should be equivalent to an analogous environmental or pharmaceutical effect.(29)

We used GWAS exposure and outcome datasets with potential sample overlap which can introduce biases with directionalities difficult to predict.(54) We used GWAS data with UKB participants removed from our outcomes where possible, but we could not remove all cases of sample overlap. Post–hoc power analyses revealed that the bipolar subtype outcomes were underpowered to detect effect sizes below an OR of 1.15 which is larger than any of our univariable effect estimates. Lastly, we restricted to participants of European ancestry and therefore we cannot necessarily generalise our results to other populations or ancestry groups. For instance, MR findings have suggested a potentially protective effect of obesity on depression in East Asian ancestry populations.(96) Further, as age data was inconsistently reported especially within our psychiatric data sources and was mainly derived during mid to late adulthood, we cannot necessarily generalise our findings to specific sensitive timing periods such as adolescence. Finally, we cannot detect non–linear effects, which warrants further investigation.

In conclusion, adult body size appears to act on multiple features of bipolar, although potential subtype–dependent effects currently remain unclear. We found stronger evidence for age–dependent effects whereby adult body size appears more detrimental to adult MDD, depressive symptoms and subthreshold mania, whereas childhood body size does not appear detrimental to adult psychiatric outcomes.

Conversely, current subtype classifications may not reflect distinct underlying mechanisms, therefore higher powered GWASs of bipolar symptoms and aetiologically defined subtypes could be used to isolate potentially opposing effects of body size across bipolar patients. Though our effect sizes were modest, future work may identify intervenable mechanisms between body size and psychiatric outcomes, targeted towards specific patient age groups or aetiologically defined subgroups for more effective treatments of mood disorders.

## Competing Interests

All other authors have no conflicts of interest to declare.

## Funders

GMP was supported by the Integrative Epidemiology Unit which receives funding from the UK Medical Research Council and the University of Bristol (MC_UU_00032/1). REW is funded by a postdoctoral fellowship from the South-Eastern Norway Regional Health Authority (2020024).

## Supporting information

STROBE-MR Checklist

## Acknowledgments

LDSC(60) and the R package ‘MR PRESSO’ version 1.0(68) analyses were carried out using the computational facilities of the <u>Advanced Computing Research Centre</u>, University of Bristol – http://www.bristol.ac.uk/acrc/. We thank all authors, including the Psychiatric Genomics Consortium (PGC), for making their GWAS datasets publicly available.

## Data Availability

Data for summary statistics is available from the respective GWAS sources outlined in Supplementary Table 1 and/or on open GWAS websites such as the ‘GWAS Catalog’ (https://www.ebi.ac.uk/gwas/) and the ‘IEU Open GWAS Project’ (https://opengwas.io/). Psychiatric GWAS sources were obtained from the Psychiatric Genomics Consortium (https://pgc.unc.edu/for-researchers/download-results/). The code to reproduce our results are available at (https://github.com/alex-monson/Age-and-Symptom-dependent-effects-between-body-size-and-bipolar).

## Notes

### Competing Interest Statement

The authors have declared no competing interest.

## References

1. Marneros A. Mood disorders: epidemiology and natural history. Psychiatry. 2006 Apr;5(4):119–22. doi:10.1383/psyt.2006.5.4.119

2. McIntyre RS, Berk M, Brietzke E, Goldstein BI, López-Jaramillo C, Kessing LV, et al. Bipolar disorders. The Lancet. 2020 Dec 5;396(10265):1841–56. doi:10.1016/S0140-6736(20)31544-0

3. American Psychiatric Association, editor. Diagnostic and statistical manual of mental disorders: DSM-5. 5th ed. Washington, D.C: American Psychiatric Association; 2013. 947 p.

4. Michalak EE, Murray G, Young AH, Lam RW. Burden of Bipolar Depression. CNS Drugs. 2008 May 1;22(5):389–406. doi:10.2165/00023210-200822050-00003

5. Elsayed OH, Ercis M, Pahwa M, Singh B. Treatment-Resistant Bipolar Depression: Therapeutic Trends, Challenges and Future Directions. NDT. 2022 Dec;Volume 18:2927–43. doi:10.2147/NDT.S273503

6. Chan JKN, Tong CHY, Wong CSM, Chen EYH, Chang WC. Life expectancy and years of potential life lost in bipolar disorder: systematic review and meta-analysis. Br J Psychiatry. 2022 Sep;221(3):567–76. doi:10.1192/bjp.2022.19

7. Fabbri C. Treatment-resistant depression: role of genetic factors in the perspective of clinical stratification and treatment personalisation. Mol Psychiatry. 2025 May;30(5):2210–8. doi:10.1038/s41380-025-02899-0

8. Zhao Z, Okusaga OO, Quevedo J, Soares JC, Teixeira AL. The potential association between obesity and bipolar disorder: A meta-analysis. Journal of Affective Disorders. 2016 Sep 15;202:120–3. doi:10.1016/j.jad.2016.05.059

9. McElroy SL, Keck PE. Obesity in Bipolar Disorder: An Overview. Curr Psychiatry Rep. 2012 Dec 1;14(6):650–8. doi:10.1007/s11920-012-0313-8

10. Vancampfort D, Vansteelandt K, Correll CU, Mitchell AJ, De Herdt A, Sienaert P, et al. Metabolic Syndrome and Metabolic Abnormalities in Bipolar Disorder: A Meta-Analysis of Prevalence Rates and Moderators. AJP. 2013 Mar;170(3):265–74. doi:10.1176/appi.ajp.2012.12050620

11. Dent R, Blackmore A, Peterson J, Habib R, Kay GP, Gervais A, et al. Changes in Body Weight and Psychotropic Drugs: A Systematic Synthesis of the Literature. PLOS ONE. 2012 Jun 15;7(6):e36889. doi:10.1371/journal.pone.0036889

12. Mansur RB, Brietzke E, McIntyre RS. Is there a “metabolic-mood syndrome”? A review of the relationship between obesity and mood disorders. Neuroscience & Biobehavioral Reviews. 2015 May 1;52:89–104. doi:10.1016/j.neubiorev.2014.12.017

13. Lawrence JM, Breunig S, Foote IF, Tallis CB, Grotzinger AD. Genomic SEM applied to explore etiological divergences in bipolar subtypes. Psychol Med. 2024 Apr;54(6):1152–9. doi:10.1017/S0033291723002957 PubMed PMID: 37885278.

14. Song J, Kuja-Halkola R, Sjölander A, Bergen SE, Larsson H, Landén M, et al. Specificity in Etiology of Subtypes of Bipolar Disorder: Evidence From a Swedish Population-Based Family Study. Biological Psychiatry. 2018 Dec 1;Bipolar Disorder: Emerging Pathophysiologic Mechanisms 84(11):810–6. doi:10.1016/j.biopsych.2017.11.014

15. Taylor V, MacQueen G. Associations between bipolar disorder and metabolic syndrome: A review. J Clin Psychiatry. 2006 Jul;67(7):1034–41. doi:10.4088/jcp.v67n0704 PubMed PMID: 16889445.

16. Kadriu B, Deng Z, Kraus C, Johnston JN, Fijtman A, Henter ID, et al. The impact of body mass index on the clinical features of bipolar disorder: A STEP-BD study. Bipolar Disorders. 2024 Mar;26(2):160–75. doi:10.1111/bdi.13370

17. Petri E, Bacci O, Barbuti M, Pacchiarotti I, Azorin J, Angst J, et al. Obesity in patients with major depression is related to bipolarity and mixed features: evidence from the BRIDGE - II - MIX study. Bipolar Disorders. 2017 Sep;19(6):458–64. doi:10.1111/bdi.12519

18. Serafini G, Gonda X, Aguglia A, Amerio A, Santi F, Pompili M, et al. Bipolar subtypes and their clinical correlates in a sample of 391 bipolar individuals. Psychiatry Research. 2019 Nov;281:112528. doi:10.1016/j.psychres.2019.112528

19. Blasco BV, García-Jiménez J, Bodoano I, Gutiérrez-Rojas L. Obesity and Depression: Its Prevalence and Influence as a Prognostic Factor: A Systematic Review. Psychiatry Investig. 2020 Aug 25;17(8):715–24. doi:10.30773/pi.2020.0099

20. Silva DA, Coutinho E da SF, Ferriani LO, Viana MC. Depression subtypes and obesity in adults: A systematic review and meta-analysis. Obesity Reviews. 2020;21(3):e12966. doi:10.1111/obr.12966

21. Reinehr T, Roth CL. Is there a causal relationship between obesity and puberty? The Lancet Child & Adolescent Health. 2019 Jan 1;3(1):44–54. doi:10.1016/S2352-4642(18)30306-7 PubMed PMID: 30446301.

22. Warnick JL, Darling KE, West CE, Jones L, Jelalian E. Weight Stigma and Mental Health in Youth: A Systematic Review and Meta-Analysis. J Pediatr Psychol. 2022 Apr 1;47(3):237–55. doi:10.1093/jpepsy/jsab110

23. Lau JYF. Developmental Aspects of Mood Disorders. In: Cowen PJ, Sharp T, Lau JYF, editors. Behavioral Neurobiology of Depression and Its Treatment [Internet]. Berlin, Heidelberg: Springer; 2013 [cited 2026 Mar 9]. p. 15–27. Available from: 10.1007/7854_2012_214 doi:10.1007/7854_2012_214

24. Pfeifer JH, Allen NB. Puberty Initiates Cascading Relationships Between Neurodevelopmental, Social, and Internalizing Processes Across Adolescence. Biological Psychiatry. 2021 Jan 15;Adolescent Brain Development and Psychopathology 89(2):99–108. doi:10.1016/j.biopsych.2020.09.002

25. Burgess S, Swanson SA, Labrecque JA. Are Mendelian randomization investigations immune from bias due to reverse causation? Eur J Epidemiol. 2021 Mar;36(3):253–7. doi:10.1007/s10654-021-00726-8

26. Lu CY. Observational studies: a review of study designs, challenges and strategies to reduce confounding. International Journal of Clinical Practice. 2009;63(5):691–7. doi:10.1111/j.1742-1241.2009.02056.x

27. Rothman KJ, Greenland S. Causation and Causal Inference in Epidemiology. Am J Public Health. 2005 Jul;95(S1):S144–50. doi:10.2105/AJPH.2004.059204

28. Uffelmann E, Huang QQ, Munung NS, De Vries J, Okada Y, Martin AR, et al. Genome-wide association studies. Nat Rev Methods Primers. 2021 Aug 26;1(1):59. doi:10.1038/s43586-021-00056-9

29. Sanderson E, Glymour MM, Holmes MV, Kang H, Morrison J, Munafò MR, et al. Mendelian randomization. Nat Rev Methods Primers. 2022 Feb 10;2(1):1–21. doi:10.1038/s43586-021-00092-5

30. Davey-Smith G. Mendelian randomization: prospects, potentials, and limitations. International Journal of Epidemiology. 2004 Feb 1;33(1):30–42. doi:10.1093/ije/dyh132

31. Sanderson E. Multivariable Mendelian Randomization and Mediation. Cold Spring Harb Perspect Med. 2021 Feb;11(2):a038984. doi:10.1101/cshperspect.a038984 PubMed PMID: 32341063; PubMed Central PMCID: PMC7849347.

32. Power GM, Palmer T, Warrington N, Heron J, Richardson TG, Didelez V, et al. A structural mean modeling Mendelian randomization approach to investigate the lifecourse effect of adiposity: applied and methodological considerations. Am J Epidemiol. 2026 Jan 8;195(1):21–31. doi:10.1093/aje/kwaf029

33. Sanderson E, Richardson TG, Morris TT, Tilling K, Davey Smith G. Estimation of causal effects of a time-varying exposure at multiple time points through multivariable mendelian randomization. PLOS Genetics. 2022 Jul 18;18(7):e1010290. doi:10.1371/journal.pgen.1010290

34. Skrivankova VW, Richmond RC, Woolf BAR, Yarmolinsky J, Davies NM, Swanson SA, et al. Strengthening the Reporting of Observational Studies in Epidemiology Using Mendelian Randomization: The STROBE-MR Statement. JAMA. 2021 Oct 26;326(16):1614–21. doi:10.1001/jama.2021.18236

35. Burgess S, Butterworth A, Thompson SG. Mendelian Randomization Analysis With Multiple Genetic Variants Using Summarized Data. Genetic Epidemiology. 2013;37(7):658–65. doi:10.1002/gepi.21758

36. Zheng J, Baird D, Borges MC, Bowden J, Hemani G, Haycock P, et al. Recent Developments in Mendelian Randomization Studies. Curr Epidemiol Rep. 2017;4(4):330–45. doi:10.1007/s40471-017-0128-6 PubMed PMID: 29226067; PubMed Central PMCID: PMC5711966.

37. Adams MJ, Streit F, Meng X, Awasthi S, Adey BN, Choi KW, et al. Trans-ancestry genome-wide study of depression identifies 697 associations implicating cell types and pharmacotherapies. Cell. 2025 Jan 14;0(0). doi:10.1016/j.cell.2024.12.002 PubMed PMID: 39814019.

38. O’Connell KS, Koromina M, Veen T van der, Boltz T, David FS, Yang JMK, et al. Genomics yields biological and phenotypic insights into bipolar disorder [Internet]. medRxiv; 2024 [cited 2025 Feb 10]. p. 2023.10.07.23296687. Available from: https://www.medrxiv.org/content/10.1101/2023.10.07.23296687v3 doi:10.1101/2023.10.07.23296687

39. Mullins N, Forstner AJ, O’Connell KS, Coombes B, Coleman JRI, Qiao Z, et al. Genome-wide association study of more than 40,000 bipolar disorder cases provides new insights into the underlying biology. Nat Genet. 2021 Jun;53(6):817–29. doi:10.1038/s41588-021-00857-4 PubMed PMID: 34002096; PubMed Central PMCID: PMC8192451.

40. Yengo L, Sidorenko J, Kemper KE, Zheng Z, Wood AR, Weedon MN, et al. Meta-analysis of genome-wide association studies for height and body mass index in ∼700000 individuals of European ancestry. Hum Mol Genet. 2018 Oct 15;27(20):3641–9. doi:10.1093/hmg/ddy271 PubMed PMID: 30124842; PubMed Central PMCID: PMC6488973.

41. Jiang L, Zheng Z, Fang H, Yang J. A generalized linear mixed model association tool for biobank-scale data. Nat Genet. 2021 Nov;53(11):1616–21. doi:10.1038/s41588-021-00954-4

42. Okbay A, Baselmans BML, De Neve JE, Turley P, Nivard MG, Fontana MA, et al. Genetic variants associated with subjective well-being, depressive symptoms, and neuroticism identified through genome-wide analyses. Nat Genet. 2016 Jun;48(6):624–33. doi:10.1038/ng.3552

43. Trubetskoy V, Pardiñas AF, Qi T, Panagiotaropoulou G, Awasthi S, Bigdeli TB, et al. Mapping genomic loci implicates genes and synaptic biology in schizophrenia. Nature. 2022 Apr;604(7906):502–8. doi:10.1038/s41586-022-04434-5

44. Richardson TG, Sanderson E, Elsworth B, Tilling K, Davey Smith G. Use of genetic variation to separate the effects of early and later life adiposity on disease risk: mendelian randomisation study. BMJ. 2020 May 6;369:m1203. doi:10.1136/bmj.m1203 PubMed PMID: 32376654.

45. Richardson TG, Mykkänen J, Pahkala K, Ala-Korpela M, Bell JA, Taylor K, et al. Evaluating the direct effects of childhood adiposity on adult systemic metabolism: a multivariable Mendelian randomization analysis. Int J Epidemiol. 2021 Mar 30;50(5):1580–92. doi:10.1093/ije/dyab051 PubMed PMID: 33783488; PubMed Central PMCID: PMC8580280.

46. Brandkvist M, Bjørngaard JH, Ødegård RA, Åsvold BO, Davey Smith G, Brumpton B, et al. Separating the genetics of childhood and adult obesity: a validation study of genetic scores for body mass index in adolescence and adulthood in the HUNT Study. Hum Mol Genet. 2020 Dec 4;29(24):3966–73. doi:10.1093/hmg/ddaa256 PubMed PMID: 33276378; PubMed Central PMCID: PMC7906755.

47. Labrecque JA, Swanson SA. Interpretation and Potential Biases of Mendelian Randomization Estimates With Time-Varying Exposures. Am J Epidemiol. 2019 Jan 1;188(1):231–8. doi:10.1093/aje/kwy204

48. Bycroft C, Freeman C, Petkova D, Band G, Elliott LT, Sharp K, et al. Genome-wide genetic data on ∼500,000 UK Biobank participants [Internet]. bioRxiv; 2017 [cited 2026 Jan 16]. p. 166298. Available from: https://www.biorxiv.org/content/10.1101/166298v1 doi:10.1101/166298

49. Locke AE, Kahali B, Berndt SI, Justice AE, Pers TH, Day FR, et al. Genetic studies of body mass index yield new insights for obesity biology. Nature. 2015 Feb 12;518(7538):197–206. doi:10.1038/nature14177 PubMed PMID: 25673413; PubMed Central PMCID: PMC4382211.

50. Bycroft C, Freeman C, Petkova D, Band G, Elliott LT, Sharp K, et al. The UK Biobank resource with deep phenotyping and genomic data. Nature. 2018;562(7726):203–9. doi:10.1038/s41586-018-0579-z PubMed PMID: 30305743; PubMed Central PMCID: PMC6786975.

51. Sudlow C, Gallacher J, Allen N, Beral V, Burton P, Danesh J, et al. UK Biobank: An Open Access Resource for Identifying the Causes of a Wide Range of Complex Diseases of Middle and Old Age. PLOS Medicine. 2015 Mar 31;12(3):e1001779. doi:10.1371/journal.pmed.1001779

52. Power GM, Tyrrell J, Frayling TM, Davey Smith G, Richardson TG. Mendelian Randomization Analyses Suggest Childhood Body Size Indirectly Influences End Points From Across the Cardiovascular Disease Spectrum Through Adult Body Size. J Am Heart Assoc. 2021 Sep 1;10(17):e021503. doi:10.1161/JAHA.121.021503 PubMed PMID: 34465205; PubMed Central PMCID: PMC8649247.

53. Howe LJ, Tudball M, Davey Smith G, Davies NM. Interpreting Mendelian-randomization estimates of the effects of categorical exposures such as disease status and educational attainment. International Journal of Epidemiology. 2022 Jun 13;51(3):948–57. doi:10.1093/ije/dyab208

54. Burgess S, Davies NM, Thompson SG. Bias due to participant overlap in two-sample Mendelian randomization. Genetic Epidemiology. 2016;40(7):597–608. doi:10.1002/gepi.21998

55. Williams CM, Poore H, Tanksley PT, Kweon H, Courchesne-Krak NS, Londono-Correa D, et al. Guidelines for Evaluating the Comparability of Down-Sampled GWAS Summary Statistics. Behav Genet. 2023 Nov;53(5–6):404–15. doi:10.1007/s10519-023-10152-z

56. Charney AW, Ruderfer DM, Stahl EA, Moran JL, Chambert K, Belliveau RA, et al. Evidence for genetic heterogeneity between clinical subtypes of bipolar disorder. Transl Psychiatry. 2017 Jan;7(1):1. doi:10.1038/tp.2016.242

57. Almeida HS, Mitjans M, Arias B, Vieta E, Ríos J, Benabarre A. Genetic differences between bipolar disorder subtypes: A systematic review focused in bipolar disorder type II. Neuroscience & Biobehavioral Reviews. 2020 Nov 1;118:623–30. doi:10.1016/j.neubiorev.2020.07.033

58. Trubetskoy V, Pardiñas AF, Qi T, Panagiotaropoulou G, Awasthi S, Bigdeli TB, et al. Mapping genomic loci implicates genes and synaptic biology in schizophrenia. Nature. 2022 Apr;604(7906):502–8. doi:10.1038/s41586-022-04434-5

59. Jiang L, Zheng Z, Qi T, Kemper KE, Wray NR, Visscher PM, et al. A resource-efficient tool for mixed model association analysis of large-scale data. Nat Genet. 2019 Dec;51(12):1749–55. doi:10.1038/s41588-019-0530-8

60. Bulik-Sullivan BK, Loh PR, Finucane HK, Ripke S, Yang J, Patterson N, et al. LD Score regression distinguishes confounding from polygenicity in genome-wide association studies. Nat Genet. 2015 Mar;47(3):291–5. doi:10.1038/ng.3211

61. Hemani G, Zheng J, Elsworth B, Wade KH, Haberland V, Baird D, et al. The MR-Base platform supports systematic causal inference across the human phenome. Loos R, editor. eLife. 2018 May 30;7:e34408. doi:10.7554/eLife.34408

62. Hartwig FP, Davies NM, Hemani G, Davey Smith G. Two-sample Mendelian randomization: avoiding the downsides of a powerful, widely applicable but potentially fallible technique. International Journal of Epidemiology. 2016 Dec 1;45(6):1717–26. doi:10.1093/ije/dyx028

63. Rees JMB, Wood AM, Burgess S. Extending the MR-Egger method for multivariable Mendelian randomization to correct for both measured and unmeasured pleiotropy. Statistics in Medicine. 2017 Dec 20;36(29):4705–18. doi:10.1002/sim.7492

64. Sanderson E, Spiller W, Bowden J. Testing and correcting for weak and pleiotropic instruments in two-sample multivariable Mendelian randomization. Stat Med. 2021 Nov 10;40(25):5434–52. doi:10.1002/sim.9133 PubMed PMID: 34338327; PubMed Central PMCID: PMC9479726.

65. Sanderson E, Spiller W, Bowden J. Testing and Correcting for Weak and Pleiotropic Instruments in Two-Sample Multivariable Mendelian Randomisation [Internet]. bioRxiv; 2020 [cited 2025 Feb 7]. p. 2020.04.02.021980. Available from: https://www.biorxiv.org/content/10.1101/2020.04.02.021980v1 doi:10.1101/2020.04.02.021980

66. Burgess S, Bowden J, Fall T, Ingelsson E, Thompson SG. Sensitivity Analyses for Robust Causal Inference from Mendelian Randomization Analyses with Multiple Genetic Variants. Epidemiology. 2017 Jan;28(1):30. doi:10.1097/EDE.0000000000000559

67. Hemani G, Tilling K, Davey Smith G. Orienting the causal relationship between imprecisely measured traits using GWAS summary data. PLOS Genetics. 2017 Nov 17;13(11):e1007081. doi:10.1371/journal.pgen.1007081

68. Verbanck M, Chen CY, Neale B, Do R. Detection of widespread horizontal pleiotropy in causal relationships inferred from Mendelian randomization between complex traits and diseases. Nat Genet. 2018 May;50(5):693–8. doi:10.1038/s41588-018-0099-7

69. R Core Team. R: A language and environment for statistical computing. R Foundation for Statistical Computing, Vienna, Austria [Internet]. 2021. Available from: URL https://www.R-project.org/.

70. Simon GE, Perlis RH. Personalized Medicine for Depression: Can We Match Patients With Treatments? AJP. 2010 Dec;167(12):1445–55. doi:10.1176/appi.ajp.2010.09111680

71. Chen W, Feng J, Jiang S, Guo J, Zhang X, Zhang X, et al. Mendelian randomization analyses identify bidirectional causal relationships of obesity with psychiatric disorders. Journal of Affective Disorders. 2023 Oct 15;339:807–14. doi:10.1016/j.jad.2023.07.044

72. Speed MS, Jefsen OH, Børglum AD, Speed D, Østergaard SD. Investigating the association between body fat and depression via Mendelian randomization. Transl Psychiatry. 2019 Aug 5;9(1):1–9. doi:10.1038/s41398-019-0516-4

73. Patsalos O, Keeler J, Schmidt U, Penninx BWJH, Young AH, Himmerich H. Diet, Obesity, and Depression: A Systematic Review. Journal of Personalized Medicine. 2021 Mar;11(3):3. doi:10.3390/jpm11030176

74. Emmer C, Bosnjak M, Mata J. The association between weight stigma and mental health: A meta-analysis. Obesity Reviews. 2020;21(1):e12935. doi:10.1111/obr.12935

75. Preiss K, Brennan L, Clarke D. A systematic review of variables associated with the relationship between obesity and depression. Obesity Reviews. 2013;14(11):906–18. doi:10.1111/obr.12052

76. Milaneschi Y, Simmons WK, van Rossum EFC, Penninx BW. Depression and obesity: evidence of shared biological mechanisms. Mol Psychiatry. 2019 Jan;24(1):18–33. doi:10.1038/s41380-018-0017-5

77. Penninx BWJH, and Lange SMM. Metabolic syndrome in psychiatric patients: overview, mechanisms, and implications. Dialogues in Clinical Neuroscience. 2018 Mar 31;20(1):63–73. doi:10.31887/DCNS.2018.20.1/bpenninx PubMed PMID: 29946213.

78. Nusslock R, Frank E. Subthreshold bipolarity: diagnostic issues and challenges. Bipolar Disord. 2011;13(7–8):587–603. doi:10.1111/j.1399-5618.2011.00957.x PubMed PMID: 22085472; PubMed Central PMCID: PMC3397420.

79. Angst J, Gamma A, Benazzi F, Ajdacic V, Eich D, Rössler W. Toward a re-definition of subthreshold bipolarity: epidemiology and proposed criteria for bipolar-II, minor bipolar disorders and hypomania. Journal of Affective Disorders. 2003 Jan 1;Validating the biopolar spectrum73(1):133–46. doi:10.1016/S0165-0327(02)00322-1

80. Lewinsohn PM, Klein DN, Seeley JR. Bipolar Disorders in a Community Sample of Older Adolescents: Prevalence, Phenomenology, Comorbidity, and Course. Journal of the American Academy of Child & Adolescent Psychiatry. 1995 Apr;34(4):454–63. doi:10.1097/00004583-199504000-00012

81. Gallagher C, Waidyatillake N, Pirkis J, Lambert K, Cassim R, Dharmage S, et al. The effects of weight change from childhood to adulthood on depression and anxiety risk in adulthood: A systematic review. Obesity Reviews. 2023;24(7):e13566. doi:10.1111/obr.13566

82. Tyrrell J, Mulugeta A, Wood AR, Zhou A, Beaumont RN, Tuke MA, et al. Using genetics to understand the causal influence of higher BMI on depression. International Journal of Epidemiology. 2019 Jun 1;48(3):834–48. doi:10.1093/ije/dyy223

83. Pathak S, Richardson TG, Sanderson E, Åsvold BO, Bhatta L, Brumpton BM. Investigating the causal effects of childhood and adulthood adiposity on later life mental health outcome: a Mendelian randomization study. BMC Med. 2025 Jan 6;23(1):4. doi:10.1186/s12916-024-03765-6

84. Richardson TG, Mykkänen J, Pahkala K, Ala-Korpela M, Bell JA, Taylor K, et al. Evaluating the direct effects of childhood adiposity on adult systemic metabolism: a multivariable Mendelian randomization analysis. International Journal of Epidemiology. 2021 Nov 10;50(5):1580–92. doi:10.1093/ije/dyab051

85. Jung SJ, Woo H taek, Cho S, Park K, Jeong S, Lee YJ, et al. Association between body size, weight change and depression: systematic review and meta-analysis. The British Journal of Psychiatry. 2017 Jul;211(1):14–21. doi:10.1192/bjp.bp.116.186726

86. Kessler RC, Berglund P, Demler O, Jin R, Merikangas KR, Walters EE. Lifetime Prevalence and Age-of-Onset Distributions of DSM-IV Disorders in the National Comorbidity Survey Replication. Arch Gen Psychiatry. 2005 Jun 1;62(6):593–602. doi:10.1001/archpsyc.62.6.593

87. Beesdo K, Höfler M, Leibenluft E, Lieb R, Bauer M, Pfennig A. Mood episodes and mood disorders: patterns of incidence and conversion in the first three decades of life. Bipolar Disorders. 2009;11(6):637–49. doi:10.1111/j.1399-5618.2009.00738.x

88. Gall SL, Sanderson K, Smith KJ, Patton G, Dwyer T, Venn A. Bi-directional associations between healthy lifestyles and mood disorders in young adults: The Childhood Determinants of Adult Health Study. Psychological Medicine. 2016 Sep;46(12):2535–48. doi:10.1017/S0033291716000738

89. Carter AR, Fraser A, Howe LD, Harris S, Hughes A. Why caution should be applied when interpreting and promoting findings from Mendelian randomisation studies. General Psychiatry. 2023 Aug 10;36(4):e101047. doi:10.1136/gpsych-2023-101047

90. Bowden J, Del Greco M. F, Minelli C, Davey Smith G, Sheehan NA, Thompson JR. Assessing the suitability of summary data for two-sample Mendelian randomization analyses using MR-Egger regression: the role of the I2 statistic. Int J Epidemiol. 2016 Sep 11;dyw220. doi:10.1093/ije/dyw220

91. Greco M FD, Minelli C, Sheehan NA, Thompson JR. Detecting pleiotropy in Mendelian randomisation studies with summary data and a continuous outcome. Statistics in Medicine. 2015 Sep 20;34(21):2926–40. doi:10.1002/sim.6522

92. Bowden J, Davey Smith G, Burgess S. Mendelian randomization with invalid instruments: effect estimation and bias detection through Egger regression. International Journal of Epidemiology. 2015 Apr 1;44(2):512–25. doi:10.1093/ije/dyv080

93. Zhao X, Yang Y, Sun BF, Zhao YL, Yang YG. FTO and Obesity: Mechanisms of Association. Curr Diab Rep. 2014 May;14(5). doi:10.1007/s11892-014-0486-0

94. Lawlor DA, Tilling K, Davey Smith G. Triangulation in aetiological epidemiology. International Journal of Epidemiology. 2016 Dec 1;45(6):1866–86. doi:10.1093/ije/dyw314

95. Munafò MR, Higgins JPT, Davey Smith G. Triangulating Evidence through the Inclusion of Genetically Informed Designs. Cold Spring Harb Perspect Med. 2021 Aug;11(8):a040659. doi:10.1101/cshperspect.a040659 PubMed PMID: 33355252; PubMed Central PMCID: PMC8327826.

96. O’Loughlin J, Casanova F, Fairhurst-Hunter Z, Hughes A, Bowden J, Watkins ER, et al. Mendelian randomisation study of body composition and depression in people of East Asian ancestry highlights potential setting-specific causality. BMC Med. 2023 Feb 1;21(1). doi:10.1186/s12916-023-02735-8

