## Supplementary material for "Symptom and Age Dependent Casual Effects of Body Size on Bipolar: A Mendelian Randomisation Study": STROBE-MR Checklist

**STROBE-MR checklist of recommended items to address in reports of Mendelian randomization studies**^1^ ^2^

| **Item No.** | **Section** | **Checklist item** | **Page No.** | **Relevant text from manuscript** |
| --- | --- | --- | --- | --- |
| 1 | **TITLE and ABSTRACT** | Indicate Mendelian randomization (MR) as the study’s design in the title and/or the abstract if that is a main purpose of the study | 1 | Title: Symptom and Age Dependent Casual Effects of Body Size on Bipolar Disorder: A Mendelian Randomisation Study |
|  | **INTRODUCTION** |  |  |  |
| 2 | **Background** | Explain the scientific background and rationale for the reported study. What is the exposure? Is a potential causal relationship between exposure and outcome plausible? Justify why MR is a helpful method to address the study question | 6  7  7-8 | “Associations with obesity are likely bidirectional with possible mechanisms including dysfunctions to hormonal and immunological systems, psychosocial factors, shared genetic liabilities and the effects of psychotropic drugs on weight gain.(11) However, current causal evidence is inconsistent, with modest effect sizes particularly in the direction of obesity on bipolar disorder.”  “Higher body size in early life may affect hormonal and metabolic processes during puberty which may affect the neurodevelopmental processes involved in the onset of bipolar disorder. Weight stigma may also increase psychosocial stresses particularly during this sensitive period leading to an increased risk when compared to adult body size measures.(21–24)”  “A challenge in epidemiology is ascertaining whether observed effects between risk factors and outcomes are causal…(28) Mendelian randomisation (MR) is a method which attempts to reduce bias from confounding by using these genetic variants as proxies for exposures to estimate their combined effects on an outcome.(29,30)” |
| 3 | **Objectives** | State specific objectives clearly, including pre-specified causal hypotheses (if any). State that MR is a method that, under specific assumptions, intends to estimate causal effects | 8  8-9 | "Mendelian randomisation (MR) is a method which attempts to reduce bias from confounding by using these genetic variants as proxies for exposures to estimate their combined effects on an outcome.(29,30) A causal effect between the exposure(s) and outcome can be inferred if various core conditions are met (outlined in Methods).”  “We then aim to account for possible heterogeneity using MR approaches by disentangling effects of body size on: 1) diagnoses of MDD and bipolar disorder (encompassing both BD–I and BD–II) 2) specific bipolar subtypes (BD–I and BD–II separately), 3) specific bipolar symptoms (subthreshold mania and depression), and 4) at different ages (childhood v. adulthood body size)… " |
|  | **METHODS** |  |  |  |
| 4 | **Study design and data sources** | Present key elements of the study design early in the article. Consider including a table listing sources of data for all phases of the study. For each data source contributing to the analysis, describe the following: | 10-15 | Summaries of the data sources are described in Methods section e.g., “Overview of data sources  All data sources used full summary statistics from mixed–sex participants of European ancestry to maintain maximum power whilst avoiding confounding through population stratification, reducing the risk of violating the second IV condition.(36) We used the largest GWASs available to maximise statistical power…”  Data sources are listed in Supplementary Table 1 |
|  | a) | Setting: Describe the study design and the underlying population, if possible. Describe the setting, locations, and relevant dates, including periods of recruitment, exposure, follow-up, and data collection, when available. | 10-15 | "All data sources use full summary statistics from mixed-sex adult participants of European ancestry”  “Adjustment for covariates differed across GWASs but included age, sex, and genetic principal components; please see the individual GWAS papers for further details (37–46) …Unless specifically stated, age was not consistently reported across data sources but GWASs recruited participants across age ranges though typically during mid to late adulthood….” |
|  | b) | Participants: Give the eligibility criteria, and the sources and methods of selection of participants. Report the sample size, and whether any power or sample size calculations were carried out prior to the main analysis | 10-11  12 | Sample sizes are reported in Supplementary Table 1 and in Data sources within Methods  "We used the largest GWASs available to maximise statistical power. Post–hoc power calculations are reported in Supplementary Table 12”  For example: “MDD. GWAS data for 357,636 MDD cases and 1,281,936 controls was obtained across 74 clinical and population samples” |
|  | c) | Describe measurement, quality control and selection of genetic variants | 15 | " Exposure SNPs were selected from full GWAS summary data based on genome–wide significance (p<5x10–8). We select independent SNPs through clumping by removing other SNPs within regions at r2 above 0.01 and within 1000 kilobases. The SNPs where then harmonised using the TwoSampleMR package version 0.6.2.(61) in which effect alleles were matched across datasets, with palindromic SNPs with similar allele frequencies (≈50%) removed.(61,62).” |
|  | d) | For each exposure, outcome, and other relevant variables, describe methods of assessment and diagnostic criteria for diseases | 10-14  12 | In the data sources subsection of Methods:  For example: “MDD. GWAS data for 357,636 MDD cases and 1,281,936 controls was obtained across 74 clinical and population samples, defining ever reporting MDD cases via medical records, clinical diagnostics and interviews, questionnaires and other self–report measures. Diagnostic criteria included the ICD–9, ICD–10, DSM–IV, or DSM-5 classifications for major depression.(37)” |
|  | e) | Provide details of ethics committee approval and participant informed consent, if relevant | N/a | Not applicable |
| 5 | **Assumptions** | Explicitly state the three core IV assumptions for the main analysis (relevance, independence and exclusion restriction) as well assumptions for any additional or sensitivity analysis | 9  17 | “The first (IV1) is the ‘relevance’ assumption, which states that the instrument Z (SNPs) must be robustly associated with the exposure. The second (IV2), is the ‘independence’ assumption, which states that the Z–Y association should be independent of genetic confounds which may be induced through population stratification, assortative mating or dynastic effects. The third assumption (IV3), ‘exclusion restriction’, states that instruments should only act on the outcome (Z–Y) through the exposure (X) (See Figure 1).(29)”  Sensitivity analyses to test the IV conditions.  “we conducted three additional sensitivity methods (MR–Egger, weighted median, weighted mode) which make different assumptions about horizontal pleiotropy.”  See the Supplementary Method for further details |
| 6 | **Statistical methods: main analysis** | Describe statistical methods and statistics used | 14-17 | See statistical analysis section & Supplementary Method  For example: “The inverse variance weighted (IVW) method was the main analysis method used to estimate the effect of the exposures on outcomes of interest in both the univariable and multivariable analyses…estimate.(35) Statistical analyses were conducted in R version 4.3.2(69). |
|  | a) | Describe how quantitative variables were handled in the analyses (i.e., scale, units, model) | 11 | Described in the data sources section:  For example: “Effect estimates can be interpreted as the change in outcome odds per 1 SD increase in liability to body size reflecting increased likelihoods of being classified into higher weight categories; from ‘thinner’ to ‘about average’ and from ‘about average’ to ‘plumper’.(52,53) Analyses were conducted using a linear regression model assuming that genetic instruments incur equal effect between each comparative weight category.(44)“ |
|  | b) | Describe how genetic variants were handled in the analyses and, if applicable, how their weights were selected | 16 | See the statistical analysis section for example: “IVW uses fixed–effect meta–analyses to take the average effect of the exposure SNPs on the outcome, weighted on the inverse variance, such that SNP estimates with smaller standard errors are given greater weighting in the overall estimate.(35) |
|  | c) | Describe the MR estimator (e.g. two-stage least squares, Wald ratio) and related statistics. Detail the included covariates and, in case of two-sample MR, whether the same covariate set was used for adjustment in the two samples | 16  10 | MR estimators are described in the statistical analysis section and in greater detail in the Supplementary Method  For instance: “The inverse variance weighted (IVW) method was the main analysis method used to estimate the effect of the exposures on outcomes of interest in both the univariable and multivariable analyses. …”  "Adjustment for covariates differed across GWASs but included age, sex, and genetic principal components; please see the individual GWAS papers for further details (37–46)… As exposure and outcome samples are derived from ancestrally homogenous populations with similar controls for population structures applied, the SNP–exposure associations are applicable to the outcome samples.” |
|  | d) | Explain how missing data were addressed | N/a | Not Applicable |
|  | e) | If applicable, indicate how multiple testing was addressed | N/a | We did not adjust for multiple testing |
| 7 | **Assessment of assumptions** | Describe any methods or prior knowledge used to assess the assumptions or justify their validity | 16-17 | Sensitivity methods are described in detail in the Supplementary Method  In main text for example:  “*Sensitivity analyses to test the IV conditions.*  The IV1 assumption (relevance) was assessed using F–statistics, or conditional F–statistics for the multivariable MR analysis.(65) To examine the robustness of the IV3 assumption (no horizontal pleiotropy), we conducted three additional sensitivity methods (MR–Egger, weighted median, weighted mode) which make different assumptions about horizontal pleiotropy…” |
| 8 | **Sensitivity analyses and additional analyses** | Describe any sensitivity analyses or additional analyses performed (e.g. comparison of effect estimates from different approaches, independent replication, bias analytic techniques, validation of instruments, simulations) | 17  14 | “Each SNP’s variance explained in the exposure and the outcome was calculated, and Steiger filtering was applied if less than 90% of the total SNPs for each exposure had a greater variance explained in the exposure than the outcome. In such cases SNPs with greater outcome variance explained were removed and the main analyses were rerun.(67) We conducted leave–one–out analyses to assess the effects of removing single SNPs on the overall IVW estimates.(29) Scatter, leave–one–out and single SNP plots were visually inspected as a sensitivity check to identify possible outliers.”  “Genetic correlations. We conducted genetic correlation analyses using linkage disequilibrium (LD) score regression (LDSC)…” |
| 9 | **Software and pre-registration** |  |  |  |
|  | a) | Name statistical software and package(s), including version and settings used | 14-17  14-15 | Described where applicable in the statistical analysis section for example:  “Univariable MR analyses were run to estimate the total effects of the genetically predicted exposures on outcomes of interest, as depicted by the Z–X–Y association in Figure 1 using the TwoSampleMR R package version 0.6.2.” |
|  | b) | State whether the study protocol and details were pre-registered (as well as when and where) | 9 | “The study protocol was not preregistered.” |
|  | **RESULTS** |  |  |  |
| 10 | **Descriptive data** |  |  |  |
|  | a) | Report the numbers of individuals at each stage of included studies and reasons for exclusion. Consider use of a flow diagram | N/a | Not applicable |
|  | b) | Report summary statistics for phenotypic exposure(s), outcome(s), and other relevant variables (e.g. means, SDs, proportions) | 11  10-14 | This information was not provided by GWASs except for adult body size: “An adult body size variable was derived using clinically measured BMI data from 453,169 UKB participants (mean age 56.5 years).”  The frequencies of cases and controls are described in the Data Sources section and Supplementary Table 1 |
|  | c) | If the data sources include meta-analyses of previous studies, provide the assessments of heterogeneity across these studies | N/a | GWAS sources did not provide this information |
|  | d) | For two-sample MR:  i.  Provide justification of the similarity of the genetic variant-exposure associations between the exposure and outcome samples  ii.  Provide information on the number of individuals who overlap between the exposure and outcome studies | 10 | “As exposure and outcome samples are derived from ancestrally homogenous populations with similar controls for population structures applied, the SNP–exposure associations are applicable to the outcome samples.”  Estimations of sample overlap are provided in Supplementary Table 11 |
| 11 | **Main results** |  |  |  |
|  | a) | Report the associations between genetic variant and exposure, and between genetic variant and outcome, preferably on an interpretable scale | 19  18 | “The relevance condition was met for all exposures, with F–statistics and conditional F–statistics (for MVMR analyses) for instruments above 10 (Supplementary Tables 1 and 2).”  R^2^ statistics are also reported in Supplementary Table 1.  To obtain variant-outcome association values:  “Reproduction of the SNP–level effects can be obtained using the analysis code provided on Github (https://github.com/alex-monson/Age-and-Symptom-dependent-effects-between-body-size-and-bipolar) using publicly available GWAS data.” |
|  | b) | Report MR estimates of the relationship between exposure and outcome, and the measures of uncertainty from the MR analysis, on an interpretable scale, such as odds ratio or relative risk per SD difference | 17-18 | Full results are reported in Supplementary Table 2  For example, “Testing for causal effects of BMI on bipolar features and MDD. In the univariable MR analysis, (IVW), adult BMI showed robust evidence for increasing the odds of MDD (OR = 1.13 95% CI [1.09, 1.16]) and subthreshold mania (OR = 1.09 95% CI [1.00, 1.19]) as well as increasing depressive symptom scores (β = 0.07 95% CI [0.05, 0.09])…” |
|  | c) | If relevant, consider translating estimates of relative risk into absolute risk for a meaningful time period | N/a | Not applicable |
|  | d) | Consider plots to visualize results (e.g. forest plot, scatterplot of associations between genetic variants and outcome versus between genetic variants and exposure) | 16 | See Figure 4 |
| 12 | **Assessment of assumptions** |  |  |  |
|  | a) | Report the assessment of the validity of the assumptions | 19 | Outlined in the Sensitivity checks section of the results. For example:  “Sensitivity checks  The relevance condition was met for all exposures, with F–statistics and conditional F–statistics (for MVMR analyses) for instruments above 10 (Supplementary Tables 1 and 2). There was no strong evidence for violation of the IV3 condition via MR Egger intercept tests, except for adult BMI on BD–I which suggested weak evidence for possible bias from horizontal pleiotropy (p = .08) (Supplementary Table 5). Leave–one–out plots indicated that removing any SNPs did not affect the conclusions of our analyses (Supplementary Figures 3b-7b)….” |
|  | b) | Report any additional statistics (e.g., assessments of heterogeneity across genetic variants, such as *I^2^*, Q statistic or E-value) | 19-20 | Reported in Supplementary Tables 4-7  In the main text: “Q statistics indicated substantial heterogeneity for all tests except for childhood body size on subthreshold mania (p = .07). Further details on the sensitivity checks, including I^2^ statistics and SIMEX corrections are reported in the Supplementary Tables 4-10.” |
| 13 | **Sensitivity analyses and additional analyses** |  |  |  |
|  | a) | Report any sensitivity analyses to assess the robustness of the main results to violations of the assumptions | 19 | Steiger filtered results are reported in Supplementary Figure 1 and Supplementary Table 9-10. MR PRESSO results are displayed in Figure 4 and in Supplementary Table 8.  In main text: “We report outlier–robust MR PRESSO estimates alongside our main results in Supplementary Table 8. The distortion test revealed substantial evidence for outliers affecting the overall effect estimates only for adult BMI on bipolar disorder (p = .03), suggesting a substantial change to the effect estimate from the removal of outliers.(68) However, this did not change our interpretations as both IVW and MR PRESSO methods predicted very weak evidence for this effect. MR PRESSO tests did indicate evidence for childhood body size affecting bipolar disorder (OR = 1.11, p = .05) and BD–I (OR = 1.15, p = 0.06) which deviate from the conclusions of the IVW and sensitivity analyses…..” |
|  | b) | Report results from other sensitivity analyses or additional analyses | 17  18 | LDSC results are displayed in Figure 3  MVMR results are reported in Figure 4 and Supplementary Table 3  For example, in main text: “MVMR analysis assessing age–dependent effects of body size. We found strong evidence suggesting that increased adult body size, after accounting for child body size, increased the odds of MDD (OR = 1.37, 95% CI [1.27 to 1.47, subthreshold mania (OR = 1.63 [1.35 to 1.98]) as well as increasing depressive symptom scores (β = 0.19 [0.14 to 0.23]), while decreasing the odds of BD–I (OR = 0.76 [0.62 to 0.93]).” |
|  | c) | Report any assessment of direction of causal relationship (e.g., bidirectional MR) | 18 | Reverse analyses are reported in Figure 4  In main text: “*Testing for the reverse effects of MDD and bipolar disorder on adult BMI.* For our bidirectional analyses, we found robust evidence that having MDD increased adult BMI (β = 0.13, 95% CI [0.08, 0.18]). We also found evidence that bipolar disorder increased adult BMI (β = 0.08, 95% CI [0.05, 0.11]) (Figure 3).” |
|  | d) | When relevant, report and compare with estimates from non-MR analyses | 17 | “Genetic correlation analyses  Adult BMI was positively correlated with adult and child body size, MDD, subthreshold mania and depressive symptom scores whilst being negatively correlated with schizophrenia (rG = -0.11 95% CI [-0.14, -0.08]) and possibly BD–I (rG = -0.06 95% CI [-0.1, -0.02]). Child body size was only negatively associated with schizophrenia (rG = –0.1, 95% CI [–0.13, –0.06]) and positively with adult body size (rG = 0.54, 95% CI [0.51, 0.58]; Figure 3). “ |
|  | e) | Consider additional plots to visualize results (e.g., leave-one-out analyses) | 19 | Scatter, leave-one-out and single-SNP forest plots of findings with strong evidence for effects are reported in the Supplementary Figures.  In main text: “Leave–one–out plots indicated that removing any SNPs did not affect the conclusions of our analyses (Supplementary Figures 3a-7a and 3b-7b). Inspection of scatter plots and single SNP forest plots indicated the presence of outliers across multiple tests (Supplementary Figures 3c-7c).” |
|  | **DISCUSSION** |  |  |  |
| 14 | **Key results** | Summarize key results with reference to study objectives | 20 | “Our study aimed to assess the effects of body size on bipolar features using a Mendelian Randomisation framework as a tool for causal inference. Due to the high heterogeneity of bipolar disorder, we explored subtype-dependent, symptom–dependent and age–dependent factors that may moderate these associations, to identify what features of bipolar disorder body size may act on and at which life stages. This may help determine individuals who are more vulnerable to specific risk factors, such as higher body size, and inform targeted interventions at relevant life stages.(70)  We found that increased adult BMI increased odds for MDD, ever having a subthreshold manic episode and depressive symptom scores but not bipolar disorder, nor its main subtypes, BD–I or BD–II. Our results replicate previous MR findings suggesting a causal effect of higher BMI on MDD.(71,72)…” |
| 15 | **Limitations** | Discuss limitations of the study, taking into account the validity of the IV assumptions, other sources of potential bias, and imprecision. Discuss both direction and magnitude of any potential bias and any efforts to address them | 24 | See limitations section of the discussion for example:  “There were limitations to our findings. For instance, I2 (reported in the Supplementary Materials) for our adult BMI and psychiatric exposures were lower than 0.9. This suggests potential violation of the ‘No Measurement Error’ (NOME) assumption which will bias MR Egger estimates towards the null unless the ‘INstrument Strength Independent of Direct Effect’ (InSIDE) assumption is also violated; in which overall directions of bias may be hard to predict.(90) Generally I2 and Q statistics were high indicating high heterogeneity and possible horizontal pleiotropy,(91) although none of our MR Egger intercept tests indicated evidence for IV3 violation, providing evidence against consistent confounding via directional horizontal pleiotropy.(92)…” |
| 16 | **Interpretation** |  |  |  |
|  | a) | Meaning: Give a cautious overall interpretation of results in the context of their limitations and in comparison with other studies | 22  23-24 | This is outlined throughout the discussion section for example:  “This disputes observational findings in the UKB that childhood body size, adjusted for adult BMI, predicts adult depression(82) but corroborates recent MVMR work by Pathak et al. (2025) finding that childhood body size independent of adult body size predicted protective effects on adult MDD.(83)…”  “Where causality is evidenced by the IVW estimate, our sensitivity analyses generally show consistent direction with comparable magnitudes, supporting robustness to bias from horizontal pleiotropy (IV3 violation). These findings strengthen confidence in our causal estimates, although triangulation with other study designs remains necessary to support stronger causal claims.(89)” |
|  | b) | Mechanism: Discuss underlying biological mechanisms that could drive a potential causal relationship between the investigated exposure and the outcome, and whether the gene-environment equivalence assumption is reasonable. Use causal language carefully, clarifying that IV estimates may provide causal effects only under certain assumptions | 20  24  8 | “Explanations linking higher body size to depression include increased social stigma(74) and decreased quality of life(75) alongside biological mechanisms like shared genetic liability and immune–metabolic pathways involving inflammation and hormone dysregulation (e.g., insulin, cortisol and leptin).(76)”  “Also, although our obesity measures include SNPs in regions that have been functionally associated with adiposity such as FTO,(93) high–powered GWASs can detect distally related genetic effects increasing pleiotropy and threatening the gene–environment equivalence assumption which states that a genetic effect on an exposure should be equivalent to an analogous environmental or pharmaceutical effect.(29)”  “A causal effect between the exposure(s) and outcome can be inferred if various core conditions are met (outlined in Methods).” |
|  | c) | Clinical relevance: Discuss whether the results have clinical or public policy relevance, and to what extent they inform effect sizes of possible interventions | 20  25 | “Due to the high heterogeneity of bipolar disorder, we explored subtype-dependent, symptom–dependent and age–dependent factors that may moderate these associations, to identify what features of bipolar disorder body size may act on and at which life stages. This may help determine individuals who are more vulnerable to specific risk factors, such as higher body size, and inform targeted interventions at relevant life stages.(70)”  “Conversely, current subtype classifications may not reflect distinct underlying mechanisms, therefore higher powered GWASs of bipolar symptoms and aetiologically defined subtypes could be used to isolate potentially opposing effects of body size across bipolar patients Though our effect sizes were modest, future work may identify intervenable mechanisms between body size and psychiatric outcomes, targeted towards specific patient age groups or aetiologically–defined subgroups for more effective treatments of mood disorders.” |
| 17 | **Generalizability** | Discuss the generalizability of the study results (a) to other populations, (b) across other exposure periods/timings, and (c) across other levels of exposure | 24-25 | “Lastly, we restricted to participants of European ancestry and therefore we cannot necessarily generalise our results to other populations or ancestry groups. For instance, MR findings have suggested a potentially protective effect of obesity on depression in East Asian ancestry populations.(96) Further, as age data was inconsistently reported especially within our psychiatric data sources and was mainly derived during mid to late adulthood, we cannot necessarily generalise our findings to specific sensitive timing periods such as adolescence. Finally, we cannot detect non–linear effects, which warrants further investigation.” |
|  | **OTHER INFORMATION** |  |  |  |
| 18 | **Funding** | Describe sources of funding and the role of funders in the present study and, if applicable, sources of funding for the databases and original study or studies on which the present study is based | 25-26 | “Funders  GMP was supported by the Integrative Epidemiology Unit which receives funding from the UK Medical Research Council and the University of Bristol (MC_UU_00032/1). REW is funded by a postdoctoral fellowship from the South-Eastern Norway Regional Health Authority (2020024).” |
| 19 | **Data and data sharing** | Provide the data used to perform all analyses or report where and how the data can be accessed, and reference these sources in the article. Provide the statistical code needed to reproduce the results in the article, or report whether the code is publicly accessible and if so, where | 26 | “Data Availability  Data for summary statistics is available from the respective GWAS sources outlined in Supplementary Table 1 and/or on open GWAS websites such as the ‘GWAS Catalog’ (https://www.ebi.ac.uk/gwas/) and the ‘IEU Open GWAS Project’ (https://opengwas.io/). The code to reproduce our results are available at (https://github.com/alex-monson/Age-and-Symptom-dependent-effects-between-body-size-and-bipolar)“ |
| 20 | **Conflicts of Interest** | All authors should declare all potential conflicts of interest | 25 | “Competing Interests  All other authors have no conflicts of interest to declare.” |

This checklist is copyrighted by the Equator Network under the Creative Commons Attribution 3.0 Unported (CC BY 3.0) license.

1.	Skrivankova VW, Richmond RC, Woolf BAR, Yarmolinsky J, Davies NM, Swanson SA, et al. Strengthening the Reporting of Observational Studies in Epidemiology using Mendelian Randomization (STROBE-MR) Statement. JAMA. 2021;under review.

2.	Skrivankova VW, Richmond RC, Woolf BAR, Davies NM, Swanson SA, VanderWeele TJ, et al. Strengthening the Reporting of Observational Studies in Epidemiology using Mendelian Randomisation (STROBE-MR): Explanation and Elaboration. BMJ. 2021;375:n2233.
