## Supplementary Material for "Symptom and Age Dependent Casual Effects of Body Size on Bipolar: A Mendelian Randomisation Study"

**Last edited:** 23/03/2026

#### Contents

Supplementary Methods Explanations and descriptions of MR methods and analyses ……………………..……4

References…………………………………………………………………………………………………….……………8

Supplementary Table 1 Summary of GWAS data sources……………………………………………………….10

Supplementary Table 2 Results of univariable MR analyses…………………………………………………...….12

Supplementary Table 3 Results of multivariable MR analyses…………………………………………………18

Supplementary Table 4 Regression Dilution $I^2$ Statistics……………………………………………………………….20

Supplementary Table 5 MR Egger Intercept Tests……………………………………………………….………22

Supplementary Table 6 SIMEX Corrections for MR Egger Estimates…………………..…………………………24

Supplementary Table 7 Q Statistics for MR Egger and IVW Tests……………………………………..…………..27

Supplementary Table 8 Full MR PRESSO Results……………………………………………………………..……..28

Supplementary Table 9 Steiger Filtering Summary………………………………………………………………..…31

Supplementary Table 10 Steiger Filtered Main Results…………………………………………………………………..33

Supplementary Table 11 Estimations of sample overlap across GWAS data sources…………………………...35

Supplementary Table 12 Post-hoc Power Calculations…………………………………………………………………36

Supplementary Figure 1 Forest Plot Comparing Steiger Filtered and non-Filtered IVW Estimates……….38

Supplementary Figure 2 Diagram Illustrating the Nature of Shared Genetic Aetiology Between Various Mental Illnesses and Their Relationships with Body Mass Index…………………...39

Supplementary Figure 3 Sensitivity Plots for the Adult BMI on MDD Analysis…………………………….........40

Supplementary Figure 4 Sensitivity Plots for the Adult BMI on Subthreshold mania Analysis……………..42

Supplementary Figure 5 Sensitivity Plots for the Adult BMI on Depressive Symptoms Analysis…………..44

Supplementary Figure 6 Sensitivity Plots for the MDD on Adult BMI Analysis……………………………….…..46

Supplementary Figure 7 Sensitivity Plots for the BD on Adult BMI Analysis……………………………………….48

### Supplementary Methods

##### Genetic Correlation Analyses

Linkage disequilibrium score regression (LDSC) estimates the genome−wide association between linkage disequilibrium (LD) and the signal strength of genetic associations with a trait or across multiple traits.(1) Single nucleotide polymorphisms (SNPs) in higher LD regions are more likely to be inherited (and thus correlated) with more surrounding SNPs increasing the probability of being in LD with SNPs causally related to a trait(s) of interest. Therefore, on average across the genome, SNPs in higher LD regions will detect greater true genetic effects, estimated by a $\chi^2$ statistic of signal strength. Crucially by regressing $\chi^2$ against LD, you can isolate true genetic effects and confounding from population stratification and cryptic relatedness. This is because such confounding will induce spurious relationships between traits and SNPs in loci uncorrelated with LD, therefore not biasing the regression slope but only influencing the intercept. LDSC can be used to estimate the genome−wide SNP heritability of a single trait or be used to estimate genetic correlations between multiple traits by instead modelling the product of signal strength (genetic covariance) across two traits as the outcome.

##### Univariable Mendelian Randomization

Exposure SNPs were selected from full GWAS summary data as being associated below the genome−wide significance threshold at $p<5x10^{-8}$. We select independent SNPs through clumping by removing other SNPs within regions at $r^2$ above 0.01 and within 1000 kilobases.

The SNPs where then harmonised using the ‘TwoSampleMR’ package in which effect alleles were matched across datasets, with palindromic SNPs with similar allele frequencies (≈50%) removed.(2,3). Plots for results were created using the R package ‘ggplot2’ version 3.5.1.(4)

*Inverse variance weighted (IVW) method.* We meta−analysed the genome−wide significant SNPs for our exposures to predict causal effects on our outcomes using the inverse variance weighted (IVW) method. IVW uses fixed−effect meta analyses to take the average effect of the exposure SNPs on the outcome, weighted on the inverse variance, such that SNP estimates with smaller standard errors are given greater weighting in the overall estimate.(5) This was the primary MR method used.

**Sensitivity Checks**

We evidence meeting the IV1 condition through reporting F−statistics for our exposures in Table 1. Instruments with F−statistics above 10 indicate evidence for meeting the IV1 condition.(6) We also conducted 3 additional sensitivity checks (MR−Egger, weighted median, weighted mode) which make different assumptions about horizontal pleiotropy (IV3). Unlike IVW, the MR Egger estimate does not fix the slope at the origin, making it robust to pleiotropy so long as the instrument−pleiotropic effects on the outcome are uncorrelated with the instrument−exposure effects; the ‘INstrument strength independent of direct effect’ assumption (InSIDE assumption). An MR Egger intercept test is also conducted where deviations of the intercept from zero (p<.05) indicate the presence of directional pleiotropy.(7) However, like with IVW, MR Egger assumes minimal measurement error in the SNP−exposure association; ‘no measurement error assumption’ (NOME). When the

NOME assumption is not met, which can be indicated if $I^2_{ZX}$ statistics are below 90%, true causal estimates can be attenuated and lead to false positive MR Egger intercept tests. In these cases, a simulation extraction (SIMEX) correction is recommended whereby increasing levels of measurement error are simulated and a corrected MR Egger estimate is predicted where measurement error equals zero.(8) SIMEX corrections were ran using the R package 'simex' version 1.8.(9) The decisions of whether to report SIMEX corrections or omit MR Egger estimates entirely are reported in Supplementary Table 3. When SIMEX corrections are applied, they are reported as the MR Egger estimates in Supplementary Table 1. We further report additional MR methods; the weighted median and weight mode methods are robust to pleiotropy in situations where at least 50% of the SNPs or the largest cluster of effects are valid predictors of the x−y association respectively.(10,11) Although IV3 violation cannot be directly observed and these tests rely on different assumptions, consistency in the direction of effects across these methods can be regarded as stronger support against IV3 violation.(12,13)

To address potential confounding effects through reverse causation we use Steiger filtering to remove SNPs where variation explained in the outcome is greater than in the exposure.(14) We also report leave−one−out analyses to assess the effects of removing single SNPs on the overall IVW estimates.(13) Scatter and single SNP plots were used to identify outliers. ~~Scatter, L~~Leave−one−out ~~and single SNP~~ plots were visually inspected as a sensitivity check to assess if single SNPs had substantial effects or biases on the overall MR estimate by sequentially removing individual effects from the analysis.(2) Outlier SNP effects caused by possible horizontal pleiotropy can be detected and removed using the MR PRESSO method (15). We report outlier corrected causal estimates in Figure 2 with full MR PRESSO results reported in ~~the~~

Supplementary Table 7 Material. This includes the outlier test results when outliers are removed, global test results which indicate the presence of horizontal pleiotropy and a distortion test indicating the extent of change in the causal estimate due to the removal of outliers.

#### References

1. Bulik-Sullivan BK, Loh PR, Finucane HK, Ripke S, Yang J, Patterson N, et al. LD Score regression distinguishes confounding from polygenicity in genome-wide association studies. Nat Genet. 2015 Mar;47(3):291–5.

2. Hartwig FP, Davies NM, Hemani G, Davey Smith G. Two-sample Mendelian randomization: avoiding the downsides of a powerful, widely applicable but potentially fallible technique. International Journal of Epidemiology. 2016 Dec 1;45(6):1717–26.

3. Hemani G, Zheng J, Elsworth B, Wade KH, Haberland V, Baird D, et al. The MR-Base platform supports systematic causal inference across the human phenome. Loos R, editor. eLife. 2018 May 30;7:e34408.

4. Wickham H. ggplot2: Elegant Graphics for Data Analysis [Internet]. Springer-Verlag New York; 2016. Available from: https://ggplot2.tidyverse.org

5. Burgess S, Butterworth A, Thompson SG. Mendelian Randomization Analysis With Multiple Genetic Variants Using Summarized Data. Genetic Epidemiology. 2013;37(7):658–65.

6. Sanderson E, Spiller W, Bowden J. Testing and Correcting for Weak and Pleiotropic Instruments in Two-Sample Multivariable Mendelian Randomisation [Internet]. bioRxiv; 2020 [cited 2025 Feb 7]. p. 2020.04.02.021980. Available from: https://www.biorxiv.org/content/10.1101/2020.04.02.021980v1

7. Bowden J, Davey Smith G, Burgess S. Mendelian randomization with invalid instruments: effect estimation and bias detection through Egger regression. International Journal of Epidemiology. 2015 Apr 1;44(2):512–25.

8. Bowden J, Del Greco M F, Minelli C, Davey Smith G, Sheehan NA, Thompson JR. Assessing the suitability of summary data for two-sample Mendelian randomization analyses using MR-Egger regression: the role of the I2 statistic. International Journal of Epidemiology. 2016 Dec 1;45(6):1961–74.

9. Lederer W, Seibold H. SIMEX- And MCSIMEX-Algorithm for Measurement Error Models [Internet]. 2019. Available from: https://CRAN.R-project.org/package=simex

10. Bowden J, Davey Smith G, Haycock PC, Burgess S. Consistent Estimation in Mendelian Randomization with Some Invalid Instruments Using a Weighted Median Estimator. Genetic Epidemiology. 2016;40(4):304–14.

11. Hartwig FP, Davey Smith G, Bowden J. Robust inference in summary data Mendelian randomization via the zero modal pleiotropy assumption. International Journal of Epidemiology. 2017 Dec 1;46(6):1985–98.

12. Burgess S, Bowden J, Fall T, Ingelsson E, Thompson SG. Sensitivity Analyses for Robust Causal Inference from Mendelian Randomization Analyses with Multiple Genetic Variants. Epidemiology. 2017 Jan;28(1):30.

13. Sanderson E, Glymour MM, Holmes MV, Kang H, Morrison J, Munafò MR, et al. Mendelian randomization. Nat Rev Methods Primers. 2022 Feb 10;2(1):1–21.

14. Hemani G, Tilling K, Smith GD. Orienting the causal relationship between imprecisely measured traits using GWAS summary data. PLOS Genetics. 2017 Nov 17;13(11):e1007081.

15. Verbanck M, Chen CY, Neale B, Do R. Detection of widespread horizontal pleiotropy in causal relationships inferred from Mendelian randomization between complex traits and diseases. Nat Genet. 2018 May;50(5):693–8.

### Supplementary Table 1

*Summary of GWAS data sources*

| Phenotype | Source | Sample Size (N) | N Cases | N Controls | Independent Genome–wide Significant SNPs | F–stat[a] | R2 %[a] |
|---|---|---|---|---|---|---|---|
| ***Exposures*** | | | | | | | |
| *Forward Direction* | | | | | | | |
| **Adult BMI** | Yengo et al. (2018)(37) | 681,275 | n/a | n/a | 1,034 | 58.3 | 8.85 |
| **Child Body Size** | Richardson et al. (2020) (38) | 453,169 | n/a | n/a | 350 | 62.4 | 4.82 |
| **Adult Body Size** | Richardson et al. (2020)(38) | 453,169 | n/a | n/a | 693 | 47.2 | 7.22 |
| *Reverse Direction* | | | | | | | |
| **Major depressive disorder (MDD) (top 10,000 independent SNPs)** | Adams et al. (2025)(39) | 3,887,532 | 525,197 | 3,362,335 | 619 | 45.1 | 0.717 |
| **Bipolar (tophits)** | O'Connell et al. (2024)(38) | 2,454,385 | 131,969 | 2,322,416 | 260 | 39.7 | 0.419 *(reported in paper all = 18.5%)* |

| | | | | | | | |
|---|---|---|---|---|---|---|---|
| | | | ***Outcomes*** | | | | |
| | | | *Psychiatric* | | | | |
| **MDD (23andme & UKB removed)** | Adams et al. (2025)(39) | 1,639,572 | 357,636 | 1,281,936 | 197 | 40.5 | 0.486 |
| **Bipolar (23andme & UKB removed)** | O'Connell et al. (2024)(38) | 780,742 | 57,833 | 722,909 | 78 | 37.9 | 0.345 |
| | | | *Symptoms and Subtypes* | | | | |
| **Bipolar type I (BD–I) (full)** | Mullins et al. (2021)(41) | 475,038 | 25,060 | 449,978 | 45 | 38.7 | 0.366 |
| **Bipolar type II (BD–II) (full)** | Mullins et al. (2021)(41) | 370,856 | 6,781 | 364,075 | 1 | 30.8 | 0.00830 |
| **Depressive Symptoms** | Okbay et al. (2016)(42) | 161,460 | n/a | n/a | 2 | 38.8 | 0.0480 |
| **Ever 'Manic'** | Jiang et al. (2019)(43) | 146,837 | 8,449 | 138,388 | 0 | n/a | n/a |
| | | | ***For Genetic Correlation Analyses Only*** | | | | |
| **Schizophrenia** | Trubetskoy et al. (2022)(44) | 127,906 | 52,017 | 75,889 | n/a | n/a | n/a |

*Note.* [a]explained by independent genome–wide significant SNPs. BMI = body mass index. SNP = single nucleotide polymorphism. MDD = major depressive disorder. UKB = United Kingdom Biobank.

### Supplementary Table 2

*Results of Univariable MR Analyses*

| Exposure | Outcome | Method | nsnp | Beta /log(odds) | Lower 95% CI | Upper 95% CI | Odds ratio | OR lower 95% CI | OR upper 95% CI | P | SE |
|---|---|---|---|---|---|---|---|---|---|---|---|
| Adult BMI | MDD | Inverse variance weighted | 931 | 0.12 | 0.09 | 0.15 | 1.13 | 1.09 | 1.16 | 1e-13 | 0.02 |
| Adult BMI | MDD | MR Egger | 931 | 0.08 | -0.04 | 0.20 | 1.08 | 0.96 | 1.22 | 0.17 | 0.06 |
| Adult BMI | MDD | Weighted median | 931 | 0.09 | 0.06 | 0.12 | 1.10 | 1.07 | 1.13 | 2e-09 | 0.02 |
| Adult BMI | MDD | Weighted mode | 931 | 0.08 | -0.02 | 0.18 | 1.09 | 0.98 | 1.20 | 0.11 | 0.05 |
| Adult BMI | Bipolar | Inverse variance weighted | 1,008 | 2e-03 | -0.06 | 0.06 | 1.00 | 0.94 | 1.06 | 0.94 | 0.03 |
| Adult BMI | Bipolar | MR Egger | 1,008 | 0.16 | -0.04 | 0.36 | 1.17 | 0.96 | 1.43 | 0.11 | 0.10 |
| Adult BMI | Bipolar | Weighted median | 1,008 | 0.02 | -0.05 | 0.09 | 1.02 | 0.95 | 1.09 | 0.56 | 0.03 |
| Adult BMI | Bipolar | Weighted mode | 1,008 | -0.04 | -0.20 | 0.13 | 0.96 | 0.82 | 1.14 | 0.67 | 0.08 |
| Adult BMI | BD–I | Inverse variance weighted | 996 | -0.06 | -0.14 | 0.03 | 0.94 | 0.87 | 1.03 | 0.19 | 0.04 |
| Adult BMI | BD–I | MR Egger | 996 | 0.19 | -0.10 | 0.48 | 1.21 | 0.90 | 1.62 | 0.20 | 0.15 |

| Exposure | Outcome | Method | nsnp | Beta /log(odds) | Lower 95% CI | Upper 95% CI | Odds ratio | OR lower 95% CI | OR upper 95% CI | P | SE |
|---|---|---|---|---|---|---|---|---|---|---|---|
| Adult BMI | BD–I | Weighted median | 996 | 3e-03 | -0.10 | 0.10 | 1.00 | 0.91 | 1.11 | 0.96 | 0.05 |
| Adult BMI | BD–I | Weighted mode | 996 | 0.07 | -0.26 | 0.40 | 1.07 | 0.77 | 1.48 | 0.67 | 0.17 |
| Adult BMI | BD–II | Inverse variance weighted | 995 | 0.05 | -0.06 | 0.16 | 1.05 | 0.94 | 1.18 | 0.36 | 0.06 |
| Adult BMI | BD–II | MR Egger | 995 | 0.08 | -0.29 | 0.45 | 1.08 | 0.75 | 1.57 | 0.69 | 0.19 |
| Adult BMI | BD–II | Weighted median | 995 | 0.08 | -0.08 | 0.24 | 1.08 | 0.92 | 1.27 | 0.34 | 0.08 |
| Adult BMI | BD–II | Weighted mode | 995 | 0.08 | -0.35 | 0.50 | 1.08 | 0.71 | 1.64 | 0.73 | 0.22 |
| Adult BMI | Subthreshold mania | Inverse variance weighted | 1,006 | 0.09 | 6e-04 | 0.17 | 1.09 | 1.00 | 1.19 | 0.05 | 0.04 |
| Adult BMI | Subthreshold mania | MR Egger | 1,006 | 0.16 | -0.13 | 0.45 | 1.17 | 0.87 | 1.57 | 0.29 | 0.15 |
| Adult BMI | Subthreshold mania | Weighted median | 1,006 | 0.10 | -0.02 | 0.22 | 1.11 | 0.98 | 1.25 | 0.10 | 0.06 |
| Adult BMI | Subthreshold mania | Weighted mode | 1,006 | 0.16 | -0.21 | 0.52 | 1.17 | 0.81 | 1.69 | 0.40 | 0.19 |
| Adult BMI | Depressive Symptoms | Inverse variance weighted | 999 | 0.07 | 0.05 | 0.09 | | | | 3e-11 | 0.01 |

| Exposure | Outcome | Method | nsnp | Beta /log(odds) | Lower 95% CI | Upper 95% CI | Odds ratio | OR lower 95% CI | OR upper 95% CI | P | SE |
|---|---|---|---|---|---|---|---|---|---|---|---|
| Adult BMI | Depressive Symptoms | MR Egger | 999 | 0.04 | -0.04 | 0.12 | | | | 0.32 | 0.04 |
| Adult BMI | Depressive Symptoms | Weighted median | 999 | 0.06 | 0.03 | 0.09 | | | | 2e-05 | 0.01 |
| Adult BMI | Depressive Symptoms | Weighted mode | 999 | 0.01 | -0.07 | 0.09 | | | | 0.78 | 0.04 |
| Child Body Size | MDD | Inverse variance weighted | 306 | -7e-03 | -0.07 | 0.06 | 0.99 | 0.93 | 1.06 | 0.83 | 0.03 |
| Child Body Size | MDD | MR Egger | 306 | 0.02 | -0.12 | 0.16 | 1.02 | 0.89 | 1.17 | 0.76 | 0.07 |
| Child Body Size | MDD | Weighted median | 306 | 0.08 | 0.02 | 0.15 | 1.09 | 1.02 | 1.16 | 0.01 | 0.03 |
| Child Body Size | MDD | Weighted mode | 306 | 0.13 | 0.04 | 0.21 | 1.13 | 1.05 | 1.23 | 3e-03 | 0.04 |
| Child Body Size | Bipolar | Inverse variance weighted | 333 | 0.02 | -0.10 | 0.14 | 1.02 | 0.91 | 1.15 | 0.70 | 0.06 |
| Child Body Size | Bipolar | MR Egger | 333 | -0.11 | -0.38 | 0.16 | 0.89 | 0.68 | 1.17 | 0.42 | 0.14 |
| Child Body Size | Bipolar | Weighted median | 333 | 0.02 | -0.12 | 0.16 | 1.02 | 0.89 | 1.18 | 0.75 | 0.07 |
| Child Body Size | Bipolar | Weighted mode | 333 | 0.01 | -0.16 | 0.18 | 1.01 | 0.85 | 1.20 | 0.88 | 0.09 |

| Exposure | Outcome | Method | nsnp | Beta /log(odds) | Lower 95% CI | Upper 95% CI | Odds ratio | OR lower 95% CI | OR upper 95% CI | P | SE |
|---|---|---|---|---|---|---|---|---|---|---|---|
| Child Body Size | BD–I | Inverse variance weighted | 336 | 0.04 | -0.13 | 0.21 | 1.04 | 0.88 | 1.23 | 0.66 | 0.09 |
| Child Body Size | BD–I | MR Egger | 336 | -0.16 | -0.55 | 0.22 | 0.85 | 0.58 | 1.25 | 0.41 | 0.20 |
| Child Body Size | BD–I | Weighted median | 336 | -0.01 | -0.22 | 0.19 | 0.99 | 0.80 | 1.21 | 0.89 | 0.10 |
| Child Body Size | BD–I | Weighted mode | 336 | -0.08 | -0.40 | 0.24 | 0.92 | 0.67 | 1.27 | 0.62 | 0.16 |
| Child Body Size | BD–II | Inverse variance weighted | 334 | -0.07 | -0.29 | 0.14 | 0.93 | 0.75 | 1.15 | 0.50 | 0.11 |
| Child Body Size | BD–II | MR Egger | 334 | -0.16 | -0.66 | 0.34 | 0.85 | 0.52 | 1.40 | 0.53 | 0.25 |
| Child Body Size | BD–II | Weighted median | 334 | -0.01 | -0.35 | 0.33 | 0.99 | 0.70 | 1.39 | 0.94 | 0.17 |
| Child Body Size | BD–II | Weighted mode | 334 | 0.03 | -0.41 | 0.46 | 1.03 | 0.66 | 1.59 | 0.91 | 0.22 |
| Child Body Size | Subthreshold mania | Inverse variance weighted | 338 | -0.13 | -0.29 | 0.03 | 0.88 | 0.75 | 1.03 | 0.11 | 0.08 |
| Child Body Size | Subthreshold mania | MR Egger | 338 | 0.03 | -0.33 | 0.39 | 1.03 | 0.72 | 1.48 | 0.88 | 0.19 |
| Child Body Size | Subthreshold mania | Weighted median | 338 | 0.09 | -0.18 | 0.37 | 1.10 | 0.84 | 1.44 | 0.50 | 0.14 |

| Exposure | Outcome | Method | nsnp | Beta /log(odds) | Lower 95% CI | Upper 95% CI | Odds ratio | OR lower 95% CI | OR upper 95% CI | P | SE |
|---|---|---|---|---|---|---|---|---|---|---|---|
| Child Body Size | Subthreshold mania | Weighted mode | 338 | 0.34 | -0.08 | 0.76 | 1.40 | 0.92 | 2.13 | 0.12 | 0.21 |
| Child Body Size | Depressive Symptoms | Inverse variance weighted | 315 | 0.02 | -0.02 | 0.07 | 1.02 | 0.98 | 1.07 | 0.28 | 0.02 |
| Child Body Size | Depressive Symptoms | MR Egger | 315 | -0.01 | -0.11 | 0.08 | 0.99 | 0.90 | 1.08 | 0.76 | 0.05 |
| Child Body Size | Depressive Symptoms | Weighted median | 315 | 0e+00 | -0.06 | 0.06 | 1.00 | 0.94 | 1.06 | 1.00 | 0.03 |
| Child Body Size | Depressive Symptoms | Weighted mode | 315 | 6e-03 | -0.07 | 0.08 | 1.01 | 0.93 | 1.09 | 0.88 | 0.04 |
| MDD | Adult BMI | Inverse variance weighted | 264 | 0.13 | 0.08 | 0.18 | | | | 2e-07 | 0.02 |
| MDD | Adult BMI | Weighted median | 264 | 0.06 | 0.03 | 0.09 | | | | 7e-06 | 0.01 |
| MDD | Adult BMI | Weighted mode | 264 | 2e-03 | -0.09 | 0.09 | | | | 0.96 | 0.05 |
| Bipolar | Adult BMI | Inverse variance weighted | 110 | 0.08 | 0.05 | 0.11 | | | | 6e-07 | 0.02 |
| Bipolar | Adult BMI | Weighted median | 110 | 0.05 | 0.03 | 0.07 | | | | 3e-05 | 0.01 |
| Bipolar | Adult BMI | Weighted mode | 110 | 0.05 | -0.01 | 0.11 | | | | 0.10 | 0.03 |

*Note.* B = beta effect size, OR =odds ratio. 95% CI indicates the upper and lower limits of 95% confidence interval of effect sizes. P = p value for each effect estimate, SE = standard error. Nsnp indicates the number of independent genome-wide significant ($p<5 \times 10^{-8}$) single nucleotide polymorphisms (snps) used in the exposure. MDD stands for major depressive disorder, BD–I and BD–II stand for bipolar type I and II respectively. BMI stands for body mass index in $kg/m^2$

### Supplementary Table 3

*Multivariable MR Results*

| Exposure | Outcome | Method | Odds Ratio / Beta | OR/β 95% CI lower | OR/β 95% CI upper | t | P | SE | Conditional F Statistic |
|---|---|---|---|---|---|---|---|---|---|
| Adjusted child body size[a] | MDD | Inverse variance weighted | 0.84 | 0.77 | 0.91 | -4.17 | 4e-05 | 0.04 | 10.63 |
| Adjusted adult body size[a] | MDD | Inverse variance weighted | 1.37 | 1.27 | 1.47 | 8.53 | 8e-17 | 0.04 | 12.66 |
| Adjusted child body size[a] | Subthreshold mania | Inverse variance weighted | 0.66 | 0.53 | 0.82 | -3.76 | 2e-04 | 0.11 | 10.83 |
| Adjusted adult body size[a] | Subthreshold mania | Inverse variance weighted | 1.63 | 1.35 | 1.98 | 5.02 | 6e-07 | 0.10 | 12.78 |
| Adjusted child body size[b] | Depressive Symptoms | Inverse variance weighted | -0.10 | -0.16 | -0.05 | -3.53 | 5e-04 | 0.03 | 10.66 |
| Adjusted adult body size[a] | Depressive Symptoms | Inverse variance weighted | 0.19 | 0.14 | 0.23 | 7.41 | 3e-13 | 0.03 | 12.31 |

| Exposure | Outcome | Method | Odds Ratio / Beta | OR/β 95% CI lower | OR/β 95% CI upper | t | P | SE | Conditional F Statistic |
|---|---|---|---|---|---|---|---|---|---|
| Adjusted child body size[a] | Bipolar disorder | Inverse variance weighted | 1.08 | 0.92 | 1.26 | 0.97 | 0.33 | 0.08 | 10.80 |
| Adjusted adult body size[a] | Bipolar disorder | Inverse variance weighted | 0.93 | 0.81 | 1.07 | -1.02 | 0.31 | 0.07 | 12.85 |
| Adjusted child body size[a] | BD–I | Inverse variance weighted | 1.21 | 0.96 | 1.52 | 1.59 | 0.11 | 0.12 | 10.87 |
| Adjusted adult body size[a] | BD–I | Inverse variance weighted | 0.76 | 0.62 | 0.93 | -2.69 | 0.007 | 0.10 | 12.78 |
| Adjusted child body size[a] | BD–II | Inverse variance weighted | 0.87 | 0.65 | 1.17 | -0.93 | 0.35 | 0.15 | 10.92 |
| Adjusted adult body size[a] | BD–II | Inverse variance weighted | 1.13 | 0.88 | 1.46 | 0.97 | 0.33 | 0.13 | 12.89 |

*Note.* The Q statistic for heterogeneity with MDD as the outcome was Q = 2683.8 (744) (p = 8.76e-217), subthreshold mania Q = 942.2 (817) (p = 0.001), depressive symptoms Q = 1022.1 (775) (p = 4.41e-9), bipolar disorder Q = 2062.5 (806) (p = 3.18e-111), BD–I Q = 1898.8 (814) (p = 2.12e-88), BD–II Q = 1023.8 (810) (p = 3.80e-7). [a]OR used as effect size measure, [b]beta used as effect size measure. MDD stands for major depressive disorder. The tests were orientated to adjusted child body size

### Supplementary Table 4

#### Regression Dilution $I^2$ Statistics

| Exposure | Outcome | $I^2$ Weighted | $I^2$ Unweighted | Decision |
|---|---|---|---|---|
| **Adult BMI** | MDD (23andme UKB removed) | 0.78 | 0.83 | Apply SIMEX correction |
| | Bipolar (23andme UKB removed) | 0.82 | 0.85 | Apply SIMEX correction |
| | BD–I | 0.82 | 0.85 | Apply SIMEX correction |
| | BD–II | 0.82 | 0.85 | Apply SIMEX correction |
| | Subthreshold mania | 0.82 | 0.85 | Apply SIMEX correction |
| | Depressive Symptoms | 0.84 | 0.85 | Apply SIMEX correction |
| **Child Body Size** | MDD (23andme UKB removed) | 0.90 | 0.92 | Use standard MR Egger |
| | Bipolar | 0.90 | 0.92 | Use standard MR Egger |
| | BD–I | 0.90 | 0.92 | Use standard MR Egger |

| | | | | |
|---|---|---|---|---|
| | BD–II | 0.90 | 0.92 | Use standard MR Egger |
| | Depressive Symptoms | 0.91 | 0.92 | Use standard MR Egger |
| | Subthreshold mania | 0.90 | 0.92 | Use standard MR Egger |
| **MDD first 10k** | Adult BMI | 0.32 | 0.47 | Do not report MR Egger |
| **Bipolar tophits** | Adult BMI | 0.0 | 0.25 | Do not report MR Egger |

Note. MDD stands for major depressive disorder, BD–I and BD–II stand for bipolar type I and II respectively. BMI stands for body mass index in $kg/m^2$.

### Supplementary Table 5

*MR Egger Intercept Tests*

| Exposure | Outcome | Egger Intercept | P | SE |
|---|---|---|---|---|
| Adult BMI | MDD (23andme and UKB removed) | 2e-04 | 0.28 | 8e-04 |
| | Bipolar (23andme and UKB removed) | -2e-03 | 0.13 | 1e-03 |
| | BD–I | -4e-03 | 0.08 | 2e-03 |
| | BD–II | -1e-03 | 0.61 | 3e-03 |
| | Subthreshold mania | -5e-04 | 0.82 | 2e-03 |
| | Depressive Symptoms | 6e-04 | 0.23 | 5e-04 |
| Child Body Size | MDD (23andme and UKB removed) | -4e-04 | 0.65 | 9e-04 |
| | Bipolar (23andme and UKB removed) | 2e-03 | 0.28 | 2e-03 |

| | | | |
|---|---|---|---|
| BD–I | 3e-03 | 0.26 | 3e-03 |
| BD–II | 1e-03 | 0.71 | 3e-03 |
| Subthreshold mania | -2e-03 | 0.33 | 2e-03 |
| Depressive Symptoms | 6e-04 | 0.39 | 6e-04 |

*Note.* SE = standard error. MDD stands for major depressive disorder, BD–I and BD–II stand for bipolar type I and II respectively. BMI stands for body mass index in $kg/m^2$.

### Supplementary Table 6

*SIMEX Corrections for MR Egger Estimates*

| Exposure | Outcome | Parameter | Model | nsnp | Estimate | t value | P value | SE |
|---|---|---|---|---|---|---|---|---|
| Adult BMI | MDD | (Intercept) | weighted | 931 | 6e-04 | 0.72 | 0.47 | 8e-04 |
| | MDD | Regression Coefficient | weighted | 931 | 0.08 | 1.38 | 0.17 | 0.06 |
| | MDD | (Intercept) | unweighted | 931 | 2e-04 | 0.21 | 0.83 | 7e-04 |
| | MDD | Regression Coefficient | unweighted | 931 | 0.12 | 2.46 | 0.01 | 0.05 |
| | Bipolar | (Intercept) | weighted | 1,008 | -2e-03 | -1.65 | 0.10 | 1e-03 |
| | Bipolar | Regression Coefficient | weighted | 1,008 | 0.16 | 1.60 | 0.11 | 0.10 |
| | Bipolar | (Intercept) | Unweighted | 1,008 | -3e-03 | -1.86 | 0.06 | 1e-03 |
| | Bipolar | Regression Coefficient | Unweighted | 1,008 | 0.16 | 1.83 | 0.07 | 0.09 |
| | BD–I | (Intercept) | weighted | 996 | -4e-03 | -1.79 | 0.07 | 2e-03 |
| | BD–I | Regression Coefficient | weighted | 996 | 0.19 | 1.29 | 0.20 | 0.15 |

| | | | | | | | | |
|---|---|---|---|---|---|---|---|---|
| | BD–I | (Intercept) | Unweighted | 996 | -4e-03 | -2.00 | 0.05 | 2e-03 |
| | BD–I | Regression Coefficient | Unweighted | 996 | 0.18 | 1.42 | 0.16 | 0.13 |
| | BD–II | (Intercept) | weighted | 995 | -5e-04 | -0.17 | 0.86 | 3e-03 |
| | BD–II | Regression Coefficient | weighted | 995 | 0.08 | 0.41 | 0.69 | 0.19 |
| | BD–II | (Intercept) | Unweighted | 995 | 6e-04 | 0.22 | 0.83 | 3e-03 |
| | BD–II | Regression Coefficient | Unweighted | 995 | 0.02 | 0.14 | 0.89 | 0.17 |
| | Subthreshold mania | (Intercept) | weighted | 1,006 | -9e-04 | -0.43 | 0.67 | 2e-03 |
| | Subthreshold mania | Regression Coefficient | weighted | 1,006 | 0.16 | 1.07 | 0.29 | 0.15 |
| | Subthreshold mania | (Intercept) | Unweighted | 1,006 | 7e-04 | 0.33 | 0.74 | 2e-03 |
| | Subthreshold mania | Regression Coefficient | Unweighted | 1,006 | 0.04 | 0.33 | 0.75 | 0.13 |
| | Depressive Symptoms | (Intercept) | weighted | 999 | 5e-04 | 0.92 | 0.36 | 5e-04 |
| | Depressive Symptoms | Regression Coefficient | weighted | 999 | 0.04 | 1.00 | 0.32 | 0.04 |

| | | | | | | | | |
|---|---|---|---|---|---|---|---|---|
| | Depressive Symptoms | (Intercept) | Unweighted | 999 | 3e-04 | 0.57 | 0.57 | 5e-04 |
| | Depressive Symptoms | Regression Coefficient | Unweighted | 999 | 0.05 | 1.58 | 0.12 | 0.03 |

*Note.* Estimate = beta effect size. SE = standard error. Nsnp indicates the number of independent genome-wide significant ($p<5 \times 10^{-8}$) single nucleotide polymorphisms (snps) used in the exposure. MDD stands for major depressive disorder, BD–I and BD–II stand for bipolar type I and II respectively. BMI stands for body mass index in $kg/m^2$.

### Supplementary Table 7

*Q Statistics for MR Egger and IVW Tests*

| ***Exposure*** | ***Outcome*** | ***Q*** | ***P*** | ***Q IVW*** | ***P*** |
|---|---|---|---|---|---|
| Adult BMI | MDD (23andme and UKB removed) | 3533.63 (929) | 5e-299 | 3538.00 (930) | 2e-299 |
| | Bipolar (23andme and UKB removed) | 2641.67 (1006) | 6e-147 | 2647.78 (1007) | 1e-147 |
| | BD–I | 2316.66 (994) | 4e-107 | 2323.68 (995) | 7e-108 |
| | BD–II | 1337.15 (993) | 1e-12 | 1337.49 (994) | 2e-12 |
| | Subthreshold mania | 1239.99 (1004) | 4e-7 | 1240.059 (1005) | 5e-7 |
| | Depressive Symptoms | 1397.64 (997) | 6e-16 | 1399.65 (998) | 5e-16 |
| Child Body Size | MDD (23andme and UKB removed) | 1168.55 (304) | 2e-101 | 1169.35 (305) | 2e-101 |
| | Bipolar (23andme and UKB removed) | 889.33 (331) | 1e-52 | 892.50 (332) | 7e-53 |
| | BD–I | 760.75 (334) | 3e-35 | 763.64 (335) | 2e-35 |
| | BD–II | 422.11 (332) | 6e-04 | 422.28 (333) | 6e-04 |
| | Subthreshold mania | 374.54 (336) | 0.07 | 375.58 (337) | 0.07 |
| | Depressive Symptoms | 422.34 (313) | 4e-05 | 423.35 (314) | 4e-05 |

MDD stands for major depressive disorder, BD–I and BD–II stand for bipolar type I and II respectively. BMI stands for body mass index in kg/$m^2$.

### Supplementary Table 8

*Full MR PRESSO Results*

| Exposure | Outcome | No. Outliers | Method | Beta | SE | T | P | Distortion Coefficient | Distortion P value | Global Coefficient | Global P value |
|---|---|---|---|---|---|---|---|---|---|---|---|
| Adult BMI | MDD | 42 | Raw | 0.12 | 0.02 | 7.44 | 2e-13 | -3.26 | 0.78 | 3546.07 | <1e-04 |
| | MDD | | Outlier-corrected | 0.12 | 0.01 | 8.78 | 8e-18 | | | | |
| | Bipolar | 29 | Raw | 4e-03 | 0.03 | 0.13 | 0.90 | 376.97 | **0.03** | 2690.94 | <1e-04 |
| | Bipolar | | Outlier-corrected | -1e-03 | 0.03 | -0.05 | 0.96 | | | | |
| | BD–I | 18 | Raw | -0.06 | 0.04 | -1.33 | 0.18 | -42.51 | 0.48 | 2359.67 | <1e-04 |
| | BD–I | | Outlier-corrected | -0.04 | 0.04 | -0.99 | 0.32 | | | | |
| | BD–II | 0 | Raw | 0.05 | 0.06 | 0.82 | 0.41 | n/a | n/a | 1372.12 | <1e-04 |
| | Subthreshold mania | 1 | Raw | 0.10 | 0.04 | 2.21 | 0.03 | -5.40 | 0.90 | 1287.33 | <1e-04 |

| Exposure | Outcome | No. Outliers | Method | Beta | SE | T | P | Distortion Coefficient | Distortion P value | Global Coefficient | Global P value |
|---|---|---|---|---|---|---|---|---|---|---|---|
| | Subthreshold mania | | Outlier-corrected | 0.10 | 0.04 | 2.36 | 0.02 | | | | |
| | Depressive Symptoms | 2 | Raw | 0.07 | 0.01 | 6.40 | 2e-10 | 0.09 | 1.00 | 1452.08 | <1e-04 |
| | Depressive Symptoms | | Outlier-corrected | 0.07 | 0.01 | 6.49 | 1e-10 | | | | |
| Child Body Size | MDD | 14 | Raw | -7e-03 | 0.03 | -0.21 | 0.83 | -119.79 | 0.36 | 1177.28 | <1e-04 |
| | MDD | | Outlier-corrected | 0.03 | 0.03 | 1.30 | 0.20 | | | | |
| | Bipolar | 13 | Raw | 0.03 | 0.06 | 0.45 | **0.65** | -71.91 | 0.49 | 908.89 | <1e-04 |
| | Bipolar | | Outlier-corrected | 0.10 | 0.05 | 1.95 | **0.05** | | | | |
| | BD–I | 10 | Raw | 0.06 | 0.09 | 0.72 | **0.47** | -56.13 | 0.53 | 785.89 | <1e-04 |
| | BD–I | 10 | Outlier-corrected | 0.14 | 0.08 | 1.88 | **0.06** | | | | |
| | BD–II | 1 | Raw | -0.07 | 0.11 | -0.63 | 0.53 | -36.31 | 0.78 | 447.46 | 4e-04 |
| | BD–II | 1 | Outlier-corrected | -0.05 | 0.11 | -0.47 | 0.64 | | | | |

| Exposure | Outcome | No. Outliers | Method | Beta | SE | T | P | Distortion Coefficient | Distortion P value | Global Coefficient | Global P value |
|---|---|---|---|---|---|---|---|---|---|---|---|
| | Subthreshold mania | 0 | Raw | -0.14 | 0.08 | -1.67 | 0.10 | n/a | n/a | 384.59 | 0.08 |
| | Depressive Symptoms | 1 | Raw | 0.02 | 0.02 | 0.98 | 0.33 | -17.98 | 0.85 | 434.06 | <1e-04 |
| | Depressive Symptoms | | Outlier-corrected | 0.03 | 0.02 | 1.22 | 0.22 | | | | |
| MDD | Adult BMI | 48 | Raw | 0.13 | 0.02 | 5.19 | 4e-07 | 7.69 | 0.40 | 3910.35 | <1e-04 |
| | Adult BMI | 48 | Outlier-corrected | 0.12 | 0.01 | 8.87 | 3e-16 | | | | |
| Bipolar | Adult BMI | 6 | Raw | 0.06 | 0.01 | 4.99 | 1e-06 | -1.48 | 0.94 | 611.54 | <1e-04 |
| | Adult BMI | | Outlier-corrected | 0.07 | 0.01 | 5.31 | 3e-07 | | | | |

Note. Method, Beta, SE, T and P columns refer to results from the MR PRESSO ran univariable MR analyses using all snps (raw) and with outliers removed (outlier corrected). P-values are based on 7,000 permutations. The Monte Carlo standard error for p-values near 0.05 is approximately ±0.0026. MDD stands for major depressive disorder, BD–I and BD–II stand for bipolar type I and II respectively. BMI stands for body mass index in $kg/m^2$.

### Supplementary Table 9

#### *Steiger Filtering Summary*

| ***Exposure*** | ***Outcome*** | ***Prevalence Exposure*** | ***Prevalence Outcome*** | ***Percentage of True SNPs %*** | **False** | **True** |
|---|---|---|---|---|---|---|
| Adult BMI | MDD (23andme UKB removed) | n/a | 0.15 | 100 | 0 | 931 |
| | Bipolar (23andme UKB removed) | n/a | 0.04 | 97.82 | 22 | 986 |
| | BD–I | n/a | 0.02 | 90.56 | 94 | 902 |
| | BD–II | n/a | 0.02 | 80.00 | 199 | 796 |
| | Subthreshold mania | n/a | 0.10 | 90.36 | 97 | 909 |
| | Depressive Symptoms | n/a | n/a | 98.60 | 14 | 985 |
| Child Body Size | MDD (23andme UKB removed) | n/a | 0.15 | 99.35 | 2 | 304 |
| | Bipolar (23andme UKB removed) | n/a | 0.04 | 93.69 | 21 | 312 |

| | | | | | | |
|---|---|---|---|---|---|---|
| | BD–I | n/a | 0.02 | 83.63 | 55 | 281 |
| | BD–II | n/a | 0.02 | 72.46 | 92 | 242 |
| | Subthreshold mania | n/a | 0.10 | 84.32 | 53 | 285 |
| | Depressive Symptoms | n/a | n/a | 95.56 | 14 | 301 |
| MDD | Adult BMI | 0.15 | n/a | 75.76 | 64 | 200 |
| Bipolar | Adult BMI | 0.04 | n/a | 94.55 | 6 | 104 |

Note. MDD stands for major depressive disorder, BD–I and BD–II stand for bipolar disorder and bipolar type I and II respectively. BMI stands for body mass index in $kg/m^2$.

### Supplementary Table 10

*Steiger Filtered Main Results*

| Exposure | Outcome | Method | nsnp | Beta | B lower 95% CI | B Upper 95% CI | odds ratio | OR lower 95% CI | OR upper 95% CI | P | se |
|---|---|---|---|---|---|---|---|---|---|---|---|
| Adult BMI | BD–II | Inverse variance weighted | 796 | 0.05 | -0.05 | 0.16 | 1.05 | 0.95 | 1.17 | 0.31 | 0.05 |
| | BD–II | MR Egger | 796 | 0.05 | -0.24 | 0.34 | 1.05 | 0.79 | 1.4 | 0.74 | 0.15 |
| | BD–II | Weighted median | 796 | 0.08 | -0.09 | 0.24 | 1.08 | 0.92 | 1.27 | 0.36 | 0.08 |
| | BD–II | Weighted mode | 796 | 0.07 | -0.38 | 0.52 | 1.07 | 0.68 | 1.68 | 0.77 | 0.23 |
| Child Body Size | BD–I | Inverse variance weighted | 281 | 0.11 | -8e-03 | 0.23 | 1.12 | 0.99 | 1.26 | 0.07 | 0.06 |
| | BD–I | MR Egger | 281 | -0.04 | -0.31 | 0.22 | 0.96 | 0.74 | 1.24 | 0.74 | 0.13 |
| | BD–I | Weighted median | 281 | -0.01 | -0.22 | 0.19 | 0.99 | 0.80 | 1.21 | 0.90 | 0.11 |
| | BD–I | Weighted mode | 281 | -0.12 | -0.48 | 0.25 | 0.89 | 0.62 | 1.28 | 0.53 | 0.19 |
| | BD–II | Inverse variance weighted | 242 | -0.02 | -0.23 | 0.20 | 0.98 | 0.79 | 1.22 | 0.88 | 0.11 |
| | BD–II | MR Egger | 242 | 0.12 | -0.34 | 0.58 | 1.13 | 0.71 | 1.79 | 0.61 | 0.24 |
| | BD–II | Weighted median | 242 | -0.01 | -0.35 | 0.33 | 0.99 | 0.70 | 1.39 | 0.94 | 0.17 |
| | BD–II | Weighted mode | 242 | 0.11 | -0.38 | 0.59 | 1.11 | 0.69 | 1.81 | 0.67 | 0.25 |

| | | | | | | | | | | | |
|---|---|---|---|---|---|---|---|---|---|---|---|
| | Subthreshold mania | Inverse variance weighted | 285 | -0.03 | -0.19 | 0.13 | 0.97 | 0.83 | 1.14 | 0.74 | 0.08 |
| | Subthreshold mania | MR Egger | 285 | 0.02 | -0.34 | 0.37 | 1.02 | 0.71 | 1.45 | 0.93 | 0.18 |
| | Subthreshold mania | Weighted median | 285 | 0.11 | -0.16 | 0.38 | 1.11 | 0.85 | 1.46 | 0.43 | 0.14 |
| | Subthreshold mania | Weighted mode | 285 | 0.38 | -0.05 | 0.81 | 1.46 | 0.95 | 2.26 | 0.09 | 0.22 |
| MDD | Adult BMI | Inverse variance weighted | 200 | 0.06 | 0.04 | 0.09 | 1.07 | 1.04 | 1.09 | 6e-07 | 0.01 |
| | Adult BMI | Weighted median | 200 | 0.04 | 0.02 | 0.07 | 1.05 | 1.02 | 1.07 | 1e-03 | 0.01 |
| | Adult BMI | Weighted mode | 200 | -6e-03 | -0.11 | 0.10 | 0.99 | 0.89 | 1.11 | 0.91 | 0.05 |

*Note.* Steiger filtering was applied to tests where the number of snps with evidence against reverse causation was below 90%. MDD stands for major depressive disorder, BD–I and BD–II stand for bipolar type I and II respectively. BMI stands for body mass index in kg/$m^2$.

### Supplementary Table 11

*Estimations of sample overlap across GWAS data sources*

| Exposure GWAS | Outcome GWAS | Predicted maximum overlap / % | Component Matching Cohorts |
|---|---|---|---|
| Adult BMI (Yengo et al., 2018) | MDD (23and me, UKB removed); Adams et al. (2025) | 6.2 | DeCODE, HUNT, NESDA |
| | Bipolar (23and me, UKB removed); O'Connell et al. (2024) | 5.9 | DeCODE, HUNT |
| | BD–I and BD–II; Mullins et al. (2021) | 14.8 | DeCODE, HUNT, UKB, wellcome Trust, |
| | Subthreshold mania; Jiang et al. (2019) | 21.6 | UKB |
| | Depressive Symptoms; Okbay et al. (2016) | 15.5 | UKB |
| | MDD | 0 | (UKB was removed) |
| | Bipolar | 0 | (UKB was removed) |
| | BD–I & BD–II | 13.2 | UKB |
| | Subthreshold mania | 32.4 | UKB |
| | Depressive Symptoms | 23.3 | UKB |
| MDD (Full) | Adult BMI | 58.9 | DeCODE, HUNT, UKB |
| Bipolar (Full) | Adult BMI | 14.6 | DeCODE, HUNT, UKB |

MDD stands for major depressive disorder, BD–I and BD–II stand for bipolar type I and II respectively. BMI stands for body mass index in $kg/m^2$.

### Supplementary Table 12

*Post-hoc Power Calculations*

Power calculations were ran on the website https://sb452.shinyapps.io/power/ for binary outcomes.

| Exposure | Outcome | Predicted Power / % | Required OR or Beta to detect an effect with 80% power |
|---|---|---|---|
| Adult BMI | MDD | 100 | 1.02 |
| | Bipolar | 2.5 | 1.05 |
| | BD–I | 24 | 1.15 |
| | BD–II | 14 | 1.18 |
| | Subthreshold mania | 50.9 | 1.13 |
| | Depressive Symptoms | 100 | 0.02 |
| Child Body Size | MDD | 21.4 | 1.03 |
| | Bipolar | 17 | 1.06 |
| | BD–I | 8.5 | 1.21 |
| | BD–II | 16 | 1.24 |
| | Subthreshold mania | 70.7 | 1.15 |
| | Depressive Symptoms | 42.2 | 0.03 |
| MDD | Adult BMI | 100 | 0.02 |

| | | | |
|---|---|---|---|
| Bipolar | Adult BMI | 100 | 0.02 |

*Note.* We run these calculations assuming a prevalence of 1% respectively for BD–I and BD–II in the underlying population. Alpha was set to 0.05. BD–I and BD–II stands for bipolar disorder type I and II respectively. BMI stands for body mass index in kg/m$^2$. Predicted power was calculated based on the odds ratios or betas (for continuous outcomes) found for the inverse variance weighted method between the exposure on the outcome.

### Supplementary Figures

#### Supplementary Figure 1

*Forest Plot Comparing Steiger Filtered and non-Filtered IVW Estimates*

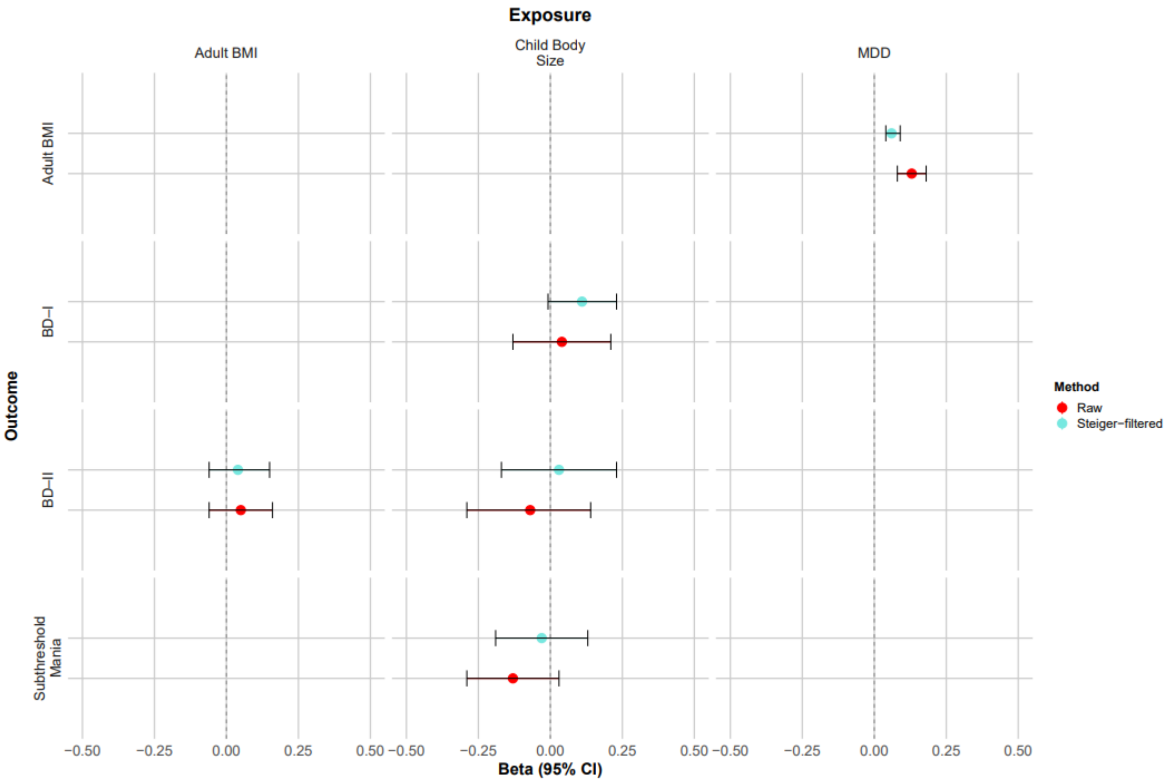


*Note.* Steiger filtering was applied to tests where the number of snps with evidence against reverse causation was below 90%. MDD stands for major depressive disorder, BD–I and BD–II stand for bipolar disorder type I and II respectively. BMI stands for body mass index in kg/m$^2$.

### Supplementary Figure 2

*Diagram Illustrating the Nature of Shared Genetic Aetiology Between Various Mental Illnesses and Their Relationships with Body Mass Index*

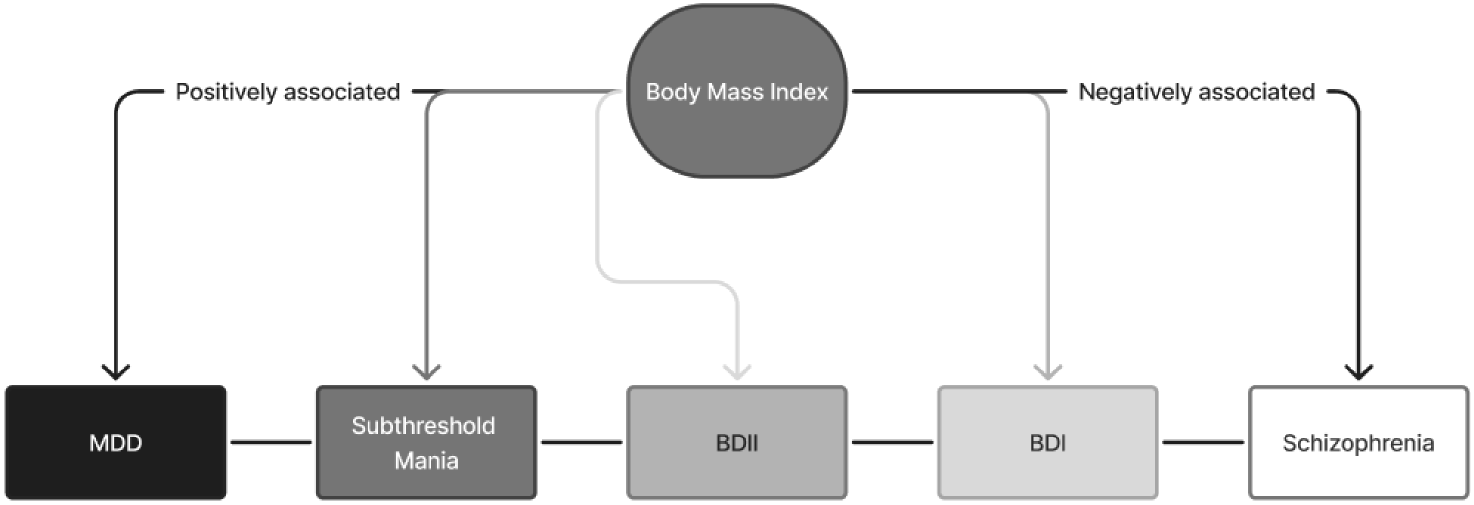


*Note.* MDD stands for major depressive disorder, BD–I with bipolar disorder type I and BD–II with bipolar disorder type II. Arrows indicate causal effects. Fainter lines indicate the tentative nature of the proposed relationship, not being evidenced by previous literature or our findings.

### Supplementary Figure 3

*Sensitivity Plots for the Adult BMI on MDD Analysis*

#### Supplementary Figure 3a

*Scatter plot of Adult BMI on MDD*

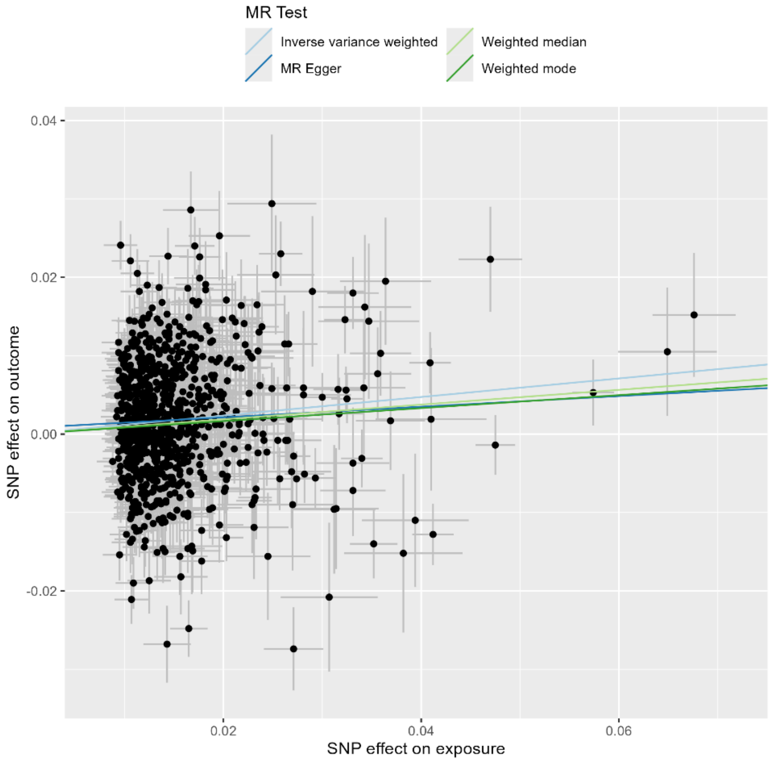

#### Supplementary Figure 3b

*Leave-one-out of Adult BMI on MDD*

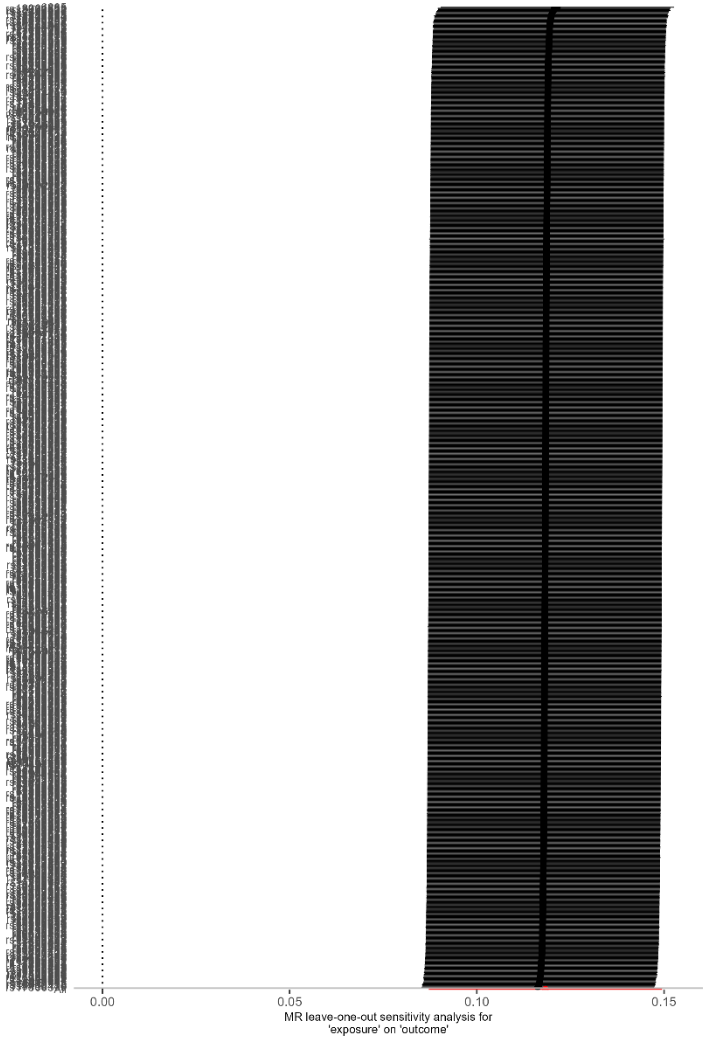


#### Supplementary Figure 3c

*Single SNP Plot of Adult BMI on MDD*

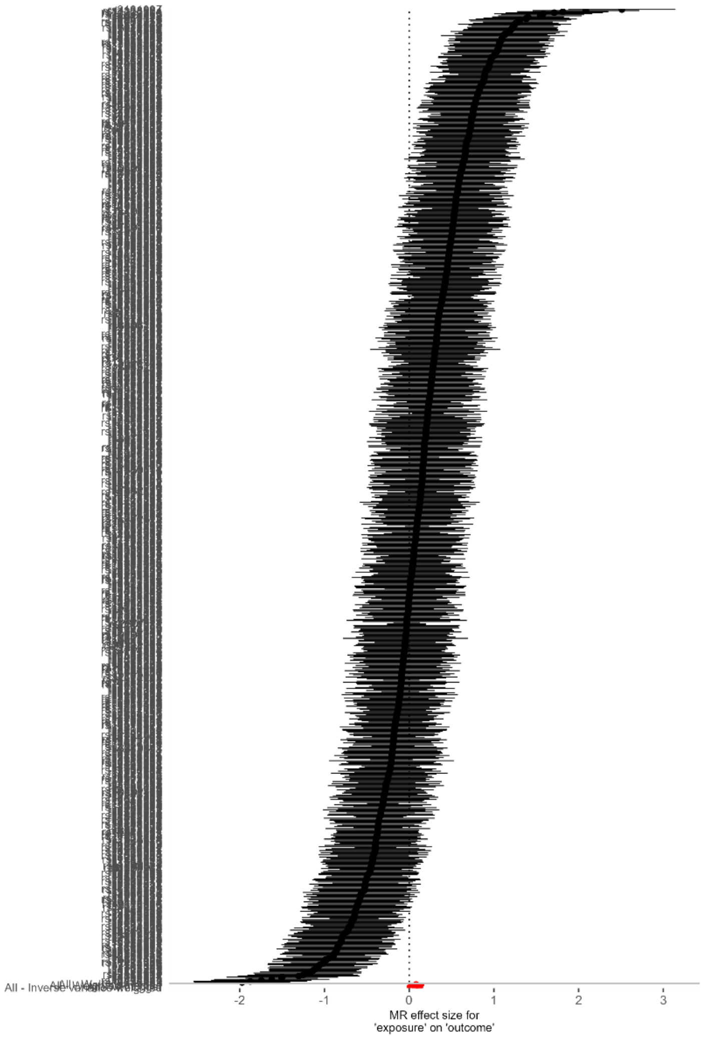

### Supplementary Table 4

*Sensitivity Plots for the Adult BMI on Subthreshold Mania Analysis*

### Supplementary Table 4a

*Scatter plot of Adult BMI on Subthreshold Mania*

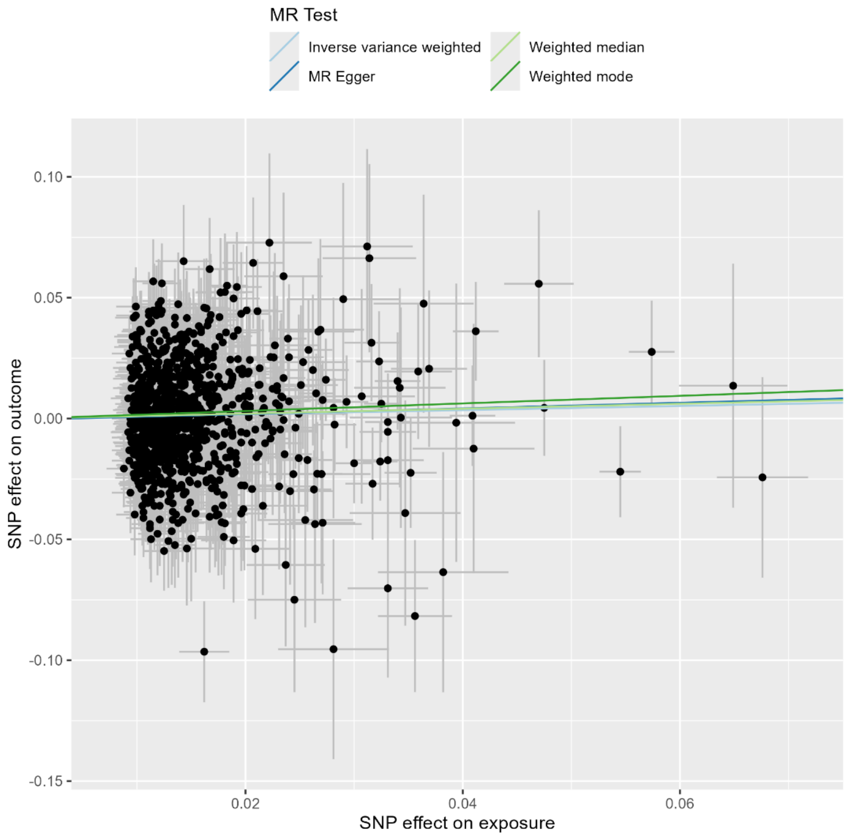

#### Supplementary Table 4b

*Leave-one-out of Adult BMI on Subthreshold Mania mania*

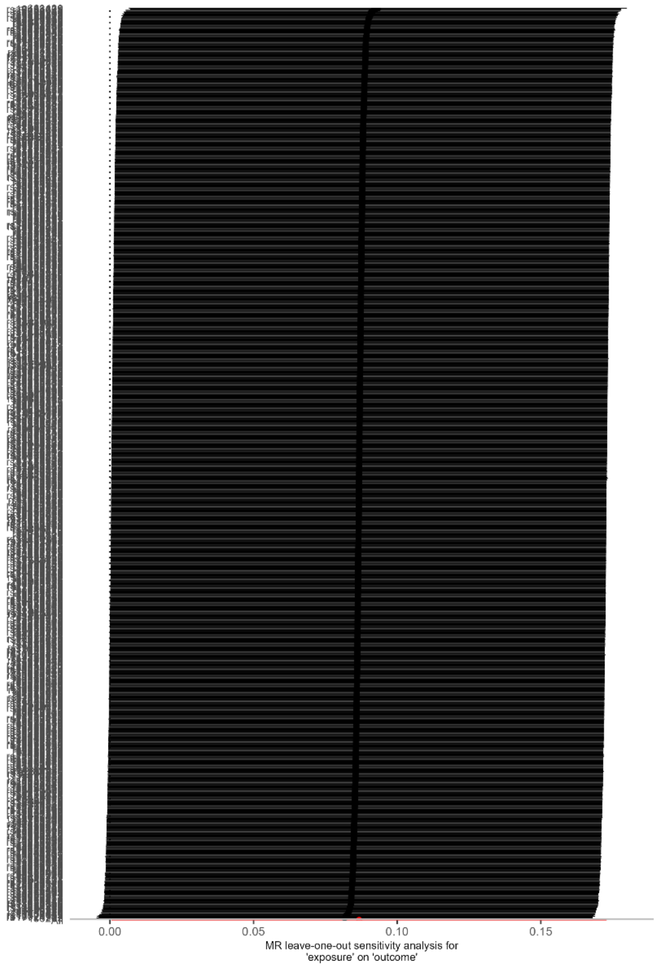


#### Supplementary Table 4c

*Single SNP Plot of Adult BMI on Subthreshold*

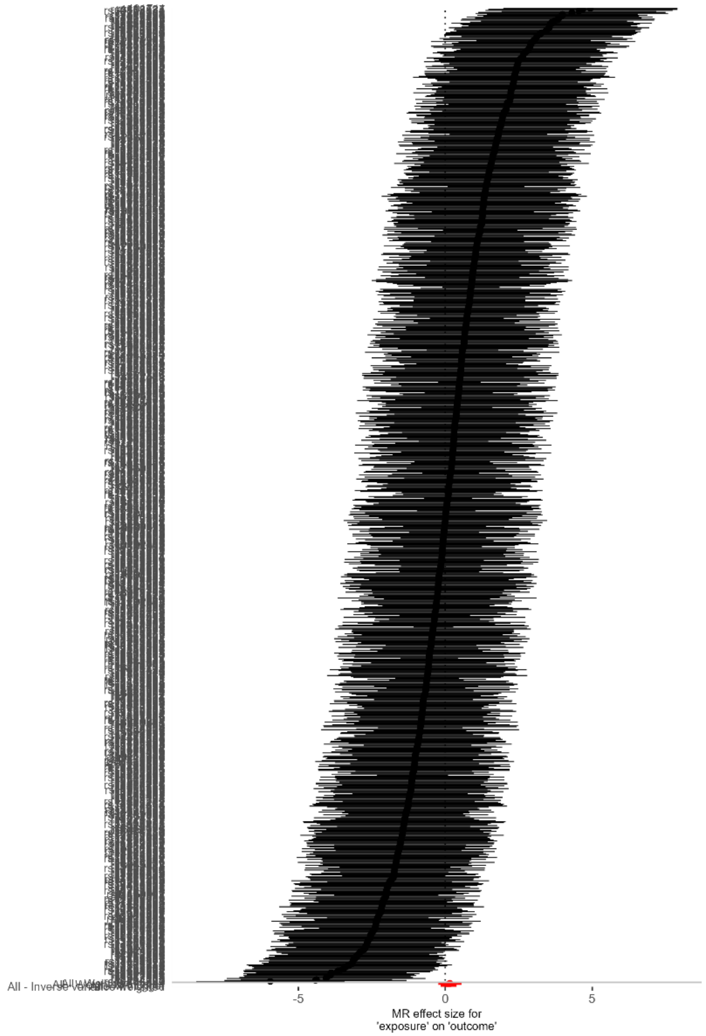

### Supplementary Table 5

*Sensitivity Plots for the Adult BMI on Depressive Symptoms*

### Supplementary Table 5a

*Scatter plot of Adult BMI on Depressive Symptoms*

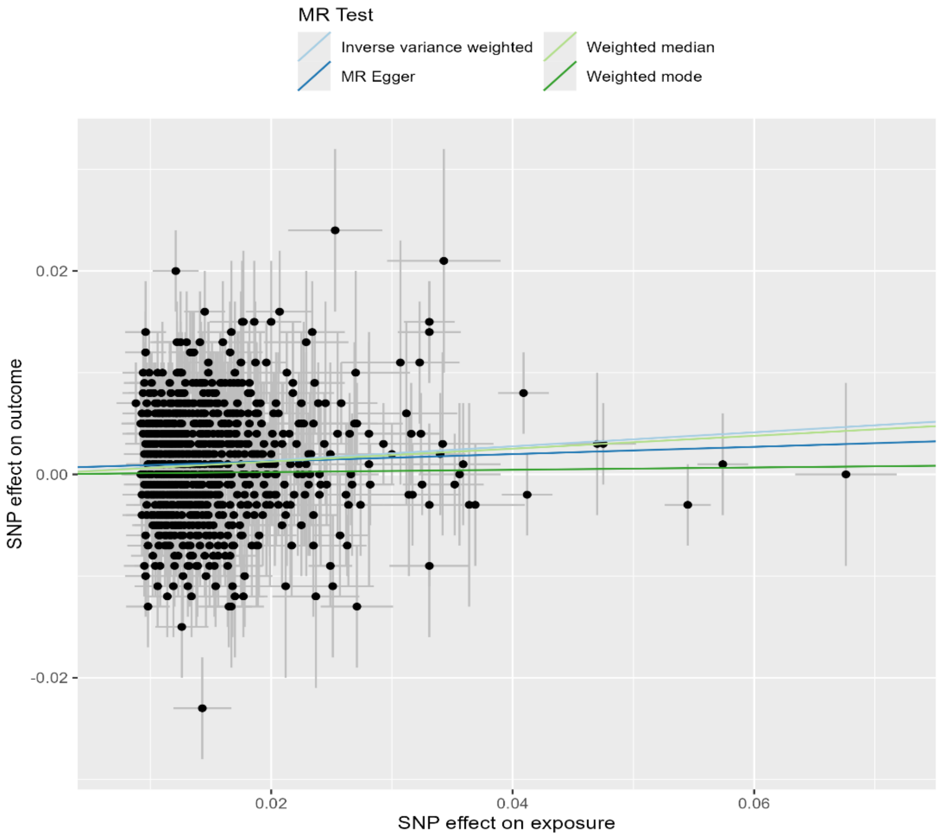

### Supplementary Table 5b

*Leave-one-out of Adult BMI on Depressive Symptoms*

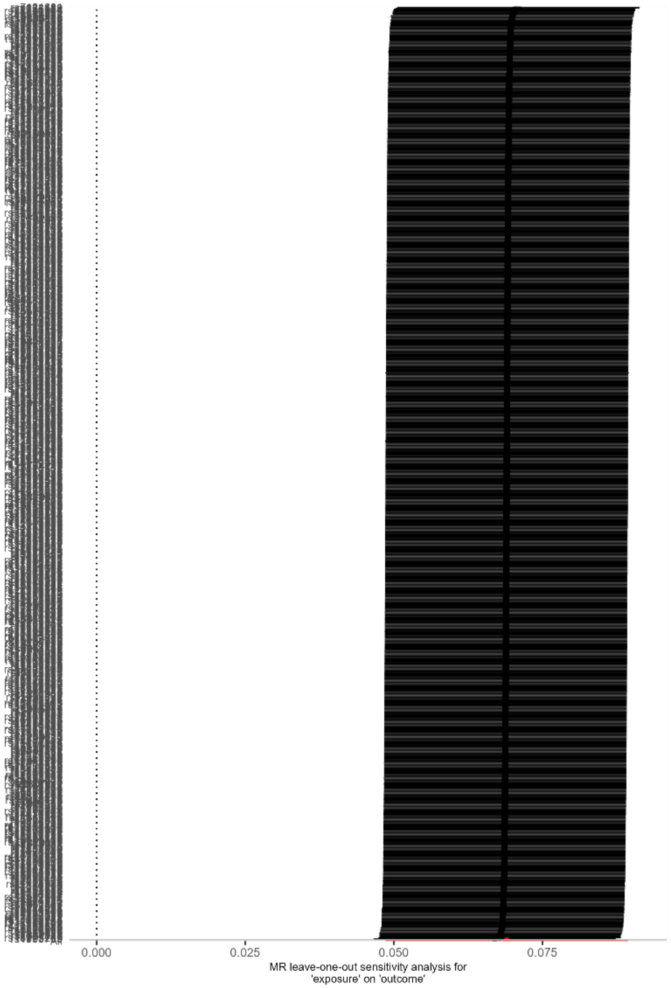


### Supplementary Table 5c

*Single SNP Plot of Adult BMI on Depressive Symptoms*

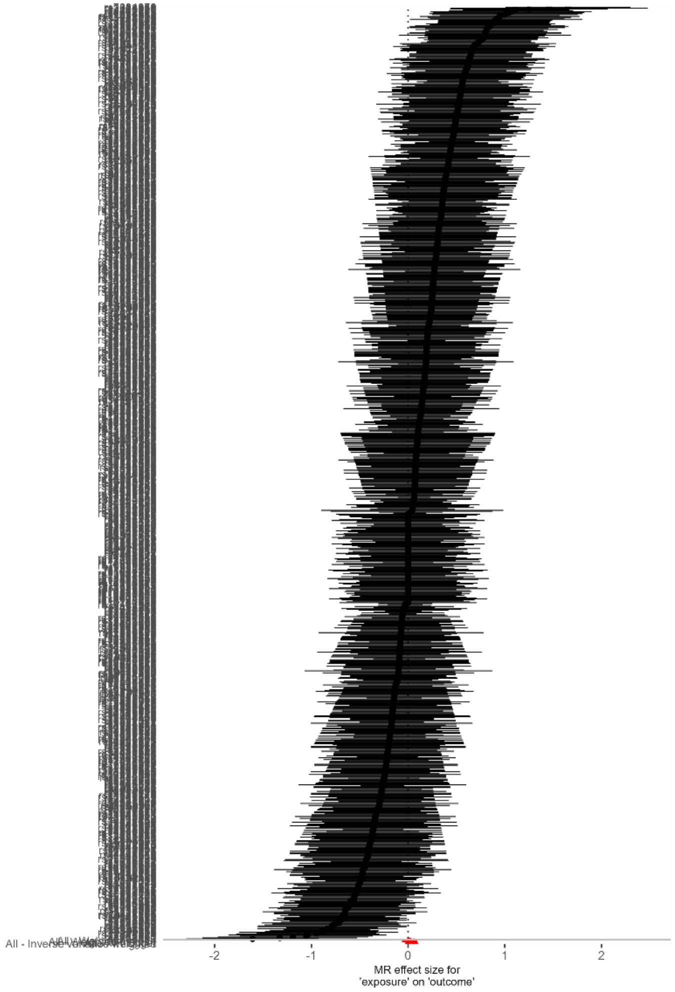

### Supplementary Table 6

*Sensitivity Plots for the MDD on Adult BMI Analysis*

### Supplementary Table 6a

*Scatter Plot of MDD on Adult BMI*

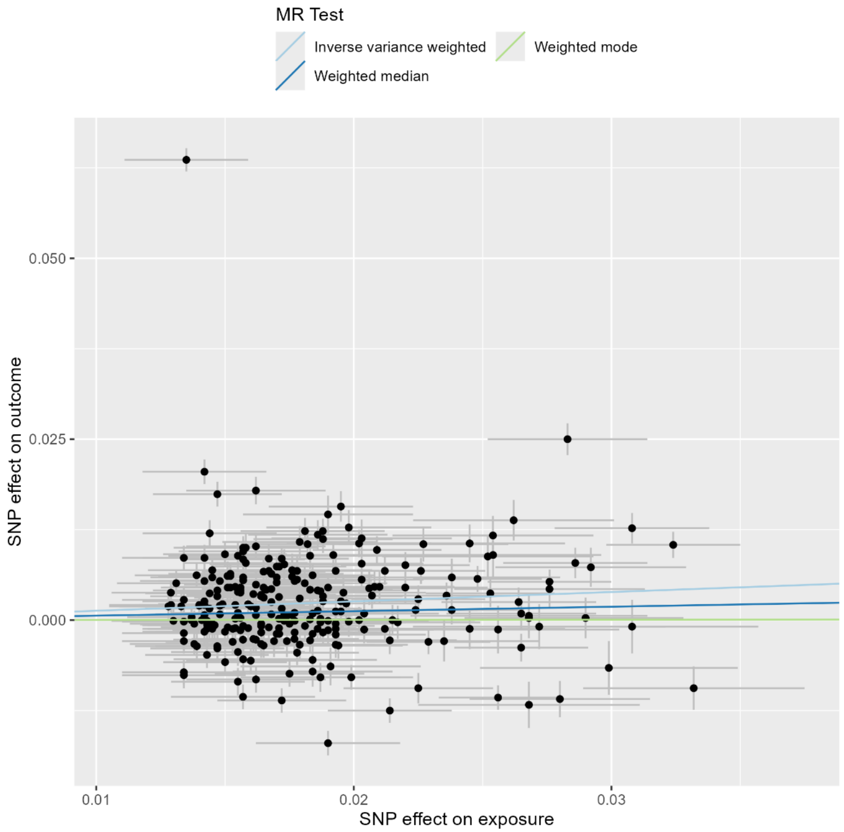

### Supplementary Table 6b

*Leave-one-out of MDD on Adult BMI*

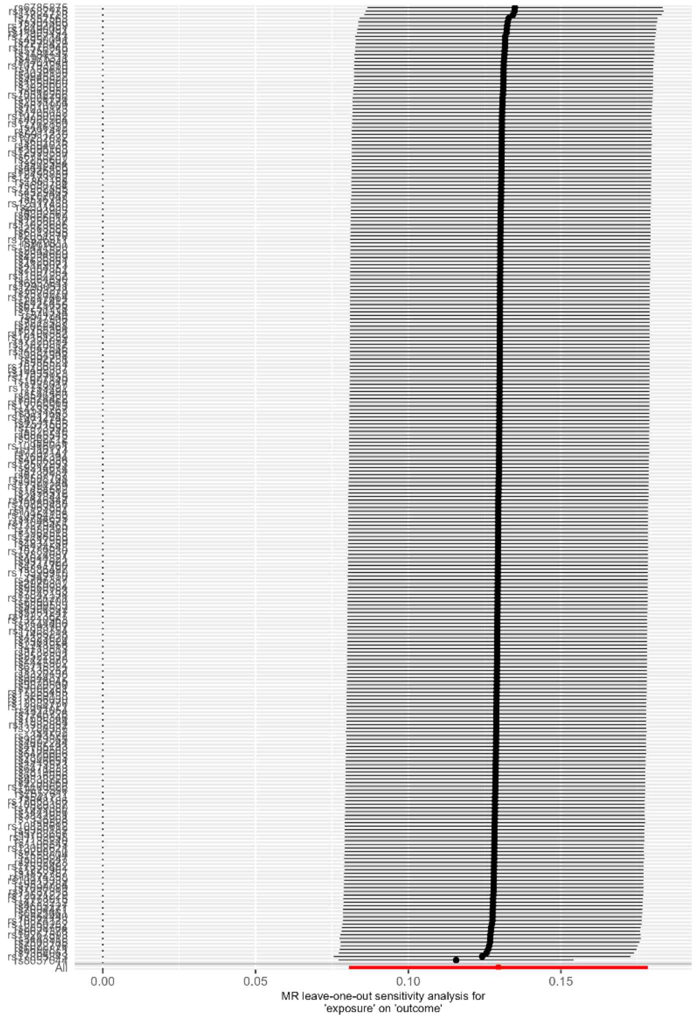


### Supplementary Table 6c

*Single SNP Plot of MDD on Adult BMI*

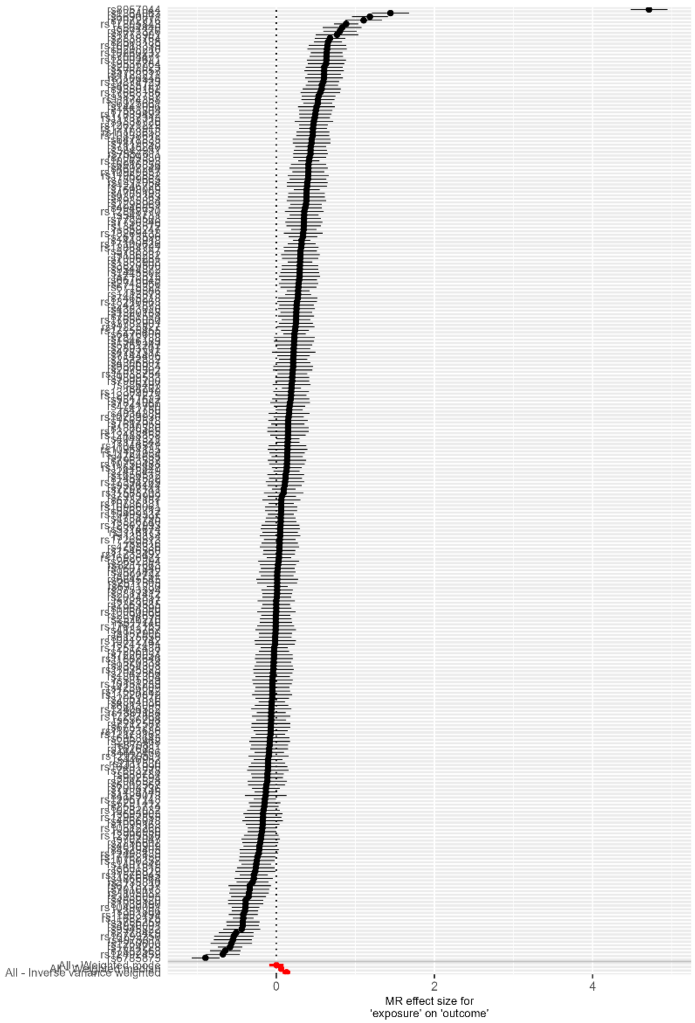

### Supplementary Table 7

*Sensitivity Plots for the Bipolar Disorder on Adult BMI Analysis*

### Supplementary Table 7a

*Scatter plot of Bipolar Disorder on Adult BMI*

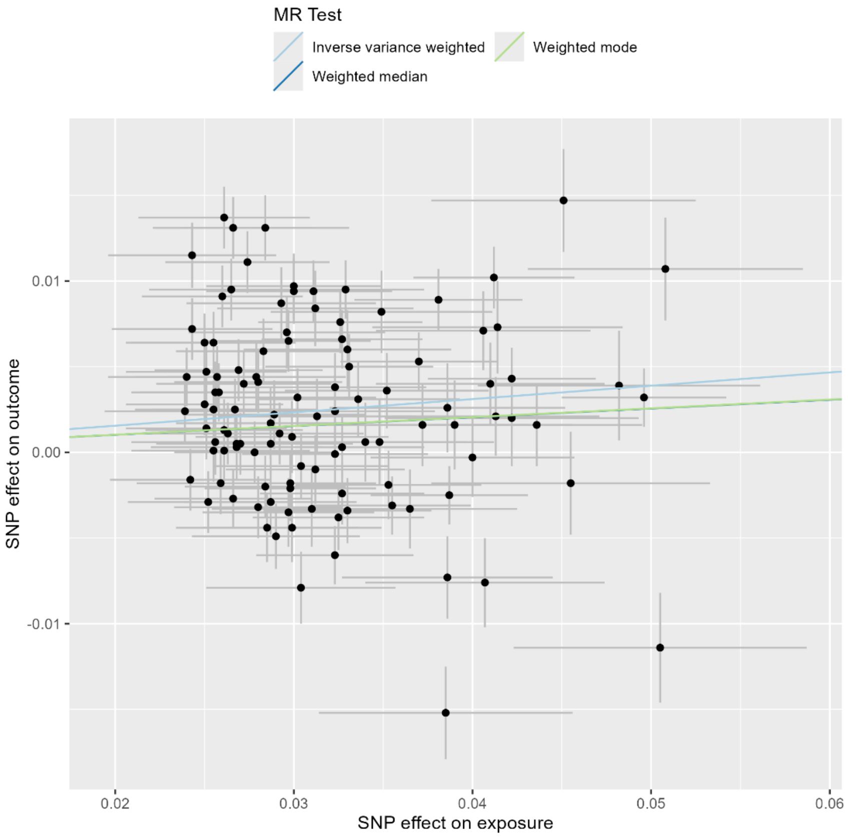

### Supplementary Table 7b

*Leave-one-out of Bipolar Disorder on Adult BMI*

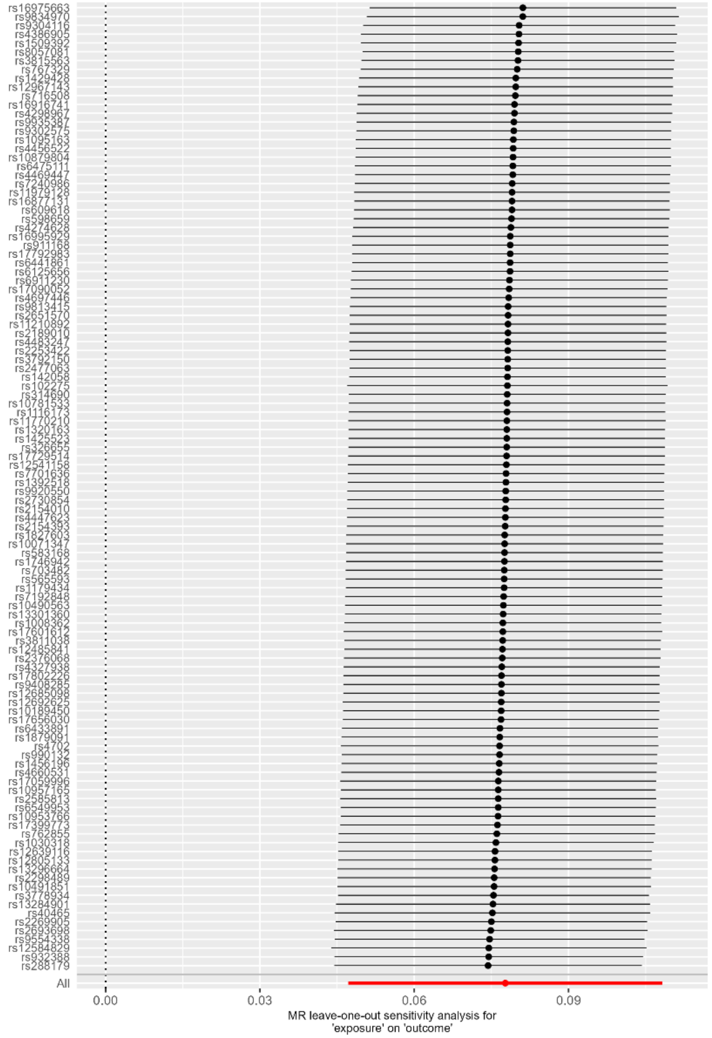


### Supplementary Table 7c

*Single SNP Plot of Bipolar Disorder on Adult BMI*

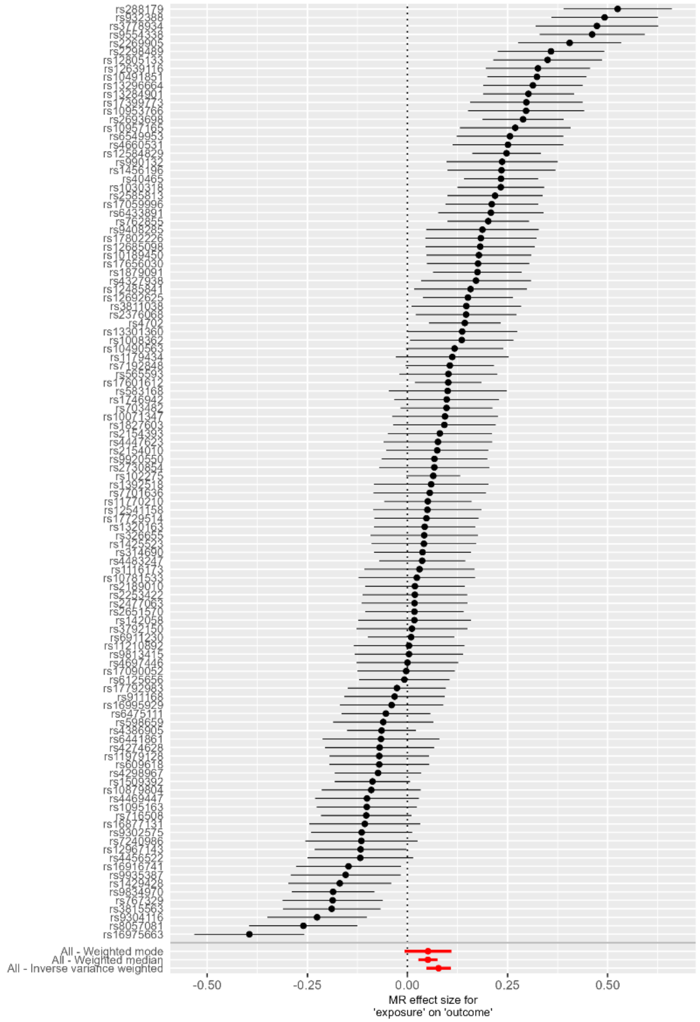
